# Dynamics of the systemic inflammatory response surrounding life events and the association with neuropsychiatric and somatic outcomes

**DOI:** 10.64898/2026.01.29.26345068

**Authors:** Rok Berlot, Ester Ipavic, Katharine Lynch-Kelly, Danish Hafeez, Timothy R. Nicholson, Mark J. Edwards, Thomas A. Pollak

## Abstract

**Background:** Adverse life events and psychosocial stressors contribute to a range of neuropsychiatric disorders. However, the role of inflammatory dynamics around stress exposure remains unclear. Using TriNetX, a large international electronic health records database, we examined how systemic inflammatory activity and its temporal dynamics relate to subsequent risk of mental illness and somatic symptoms.

**Methods:** We compared 36,772 individuals with records of adverse life events and leukocytosis in the surrounding period with matched individuals with normal leukocyte counts, and performed an analogous comparison for socioeconomic and psychosocial stressors in cohorts of 87,936 individuals with leukocytosis and matched controls. To contrast dynamic with static inflammatory responses, we compared cohorts exhibiting leukocyte count changes with those maintaining persistently normal or persistently elevated leukocyte counts around stressor exposure. Outcomes, including new mental health and somatic symptom presentations, were evaluated within two years of the stressor.

**Results:** Following acute stressors, leukocytosis (compared with normal leukocyte counts) was associated with lower rates of subsequent anxiety disorders, cognitive symptoms and several somatic symptom diagnoses, with similar reductions in anxiety observed after chronic stressors. A dynamic inflammatory response was associated with the most favourable outcomes, with lower rates of anxiety, depression, functional neurological disorder, cognitive and sleep difficulties, fatigue, and multiple pain-related and other somatic symptoms than persistently low or high inflammation.

**Conclusion:** Our findings suggest that a well-regulated inflammatory response to stressors is associated with a reduced risk of diverse mental health and somatic outcomes, and that a transient immune activation may support recovery rather than confer risk.

## 1. Introduction

Adverse life events and psychosocial stressors are major risk factors for mental illness and contribute to a broad spectrum of neurological and medical conditions, including autoimmune, neuroinflammatory, and neurodegenerative diseases (Gillespie et al., 2024; Hogg et al., 2023; Jesuthasan et al., 2025; Vaidya et al., 2024). Historically, stressors and adverse life events were also considered primary causes of a variety of symptoms occurring in the context of so-called ‘functional’ diagnoses, including functional neurological disorder (FND) and functional gastrointestinal disorders, among others (Berlot et al., 2025; Drossman, 2016). Contemporary approaches recognise that these symptoms may occur within a variety of diagnoses, which can be broadly categorised as ‘interface disorders’ (to denote their occurrence at the ‘interface’ between general medicine, neurology and/or psychiatry) (Maggio et al., 2024), or in isolation. Most current models instead situate stressors alongside other risk factors within a broader biopsychosocial framework involving multiple interacting mechanisms (Keynejad et al., 2019). Despite this shift, the biological pathways linking stressors to the triggering or exacerbation of these diverse conditions remain poorly understood. Stress-sensitive dysregulation of bodily systems has been implicated (von Majewski et al., 2023; Weber et al., 2023), and while the relevant biological mediators are unclear, systemic inflammation – particularly in the period surrounding stressor exposure – may play an important role.

The available evidence largely indicates that inflammation acts as a harmful mediator of pathology, with studies suggesting an association between a systemic and, in some cases, central nervous system-restricted pro-inflammatory state and the emergence of several neuropsychiatric disorders (Dowlati et al., 2010; Montoya et al., 2017; Paredes-Echeverri et al., 2022). Similarly, various types of stressors have been associated with prolonged increases in systemic inflammatory markers (Baldwin et al., 2018; Danese et al., 2008; Entringer et al., 2020; Yang et al., 2014). This pro-inflammatory profile, typically reflected in altered leukocyte counts, C-reactive protein or circulating cytokines, represents a reproducible biomarker pattern of the stressor response, but does not necessarily imply a direct pro-inflammatory mechanism or immunological dysfunction (Khandaker et al., 2017). While inflammation has been suggested as a key pathway by which adverse experiences become biologically embedded, the contribution of the acute inflammatory response to long-term outcomes remains to be fully elucidated. Recent findings from human and animal studies paint a more complex picture, in which acute inflammatory responses to stressors may exert adaptive or protective effects. From this viewpoint, inflammation is indeed an immunological defence mechanism, but may more fundamentally act as a mediator of tissue repair, remodelling, and homeostatic recalibration in response to stress or damage (Meizlish et al., 2021). Such an understanding may facilitate the incorporation of findings that, on a simplistic ‘defence mechanism’ framing, are hard to reconcile. For example, lower levels of proinflammatory cytokines immediately after a traumatic experience have been associated with an increased risk of post-traumatic stress disorder (PTSD) (Michopoulos et al., 2020), suggesting that a deficient neuroimmune response to acute trauma might be a mechanism underlying the development of PTSD (Heim, 2020). Evidence from genetic studies further challenges a simple deleterious model of inflammation. While elevated C-reactive protein (CRP) levels have been associated with increased risk of several mental health outcomes, individuals with genetically elevated CRP levels show a lower risk, suggesting important heterogeneity in inflammatory profiles (Milaneschi et al., 2021).

Much of the literature on inflammation and mental health has relied on a static, ‘high versus low’ approach to inflammatory markers, which obscures the dynamic nature of immune responses to psychological (or indeed any other) stressors (Marsland et al., 2017). Such framing tends to overlook the adaptive physiological functions of inflammation (Medzhitov, 2008), casting it in the rather restrictive light of a toxic risk factor. From this perspective, mild inflammatory responses may be especially informative, as they may reflect adaptive immune recalibration rather than deleterious immune activation. Further, it has previously been proposed that disease trajectories are not dependent on the level of inflammation itself, but on the initiation, regulation, and resolution of the inflammatory response (Ahuja et al., 2023). A dynamic inflammatory response to stressors could therefore facilitate neuroimmune interactions to help restore homeostasis. In contrast, persistent inflammation or a blunted immune response may fail to engage such reparative mechanisms.

In this study, we used electronic health records (EHR) to explore how inflammatory activity in the period surrounding documented stressors relates to the subsequent risk of diagnoses of mental health disorders, common somatic symptoms, and interface disorders. We focused on records of acute stressors, such as adverse life events, as well as chronic stressors, including psychosocial and socioeconomic difficulties. We compared outcomes between: i) individuals with versus without markers of mild systemic inflammation; and ii) individuals exhibiting a transiently elevated inflammatory response in close temporal proximity to adversity versus those with either a) no evidence of systemic inflammation or b) evidence of sustained inflammatory activity in that period.

## 2. Methods

### 2.1. Data and study design

The study utilised TriNetX, a global federated health research network comprising de-identified EHR data from over 168 million patients. The database contains structured clinical data, including demographic information, diagnoses coded with ICD-10-CM (International Classification of Diseases, Tenth Revision, Clinical Modification), and laboratory results. The data is updated continuously. Contributing healthcare organisations include hospitals, specialist providers, and primary care facilities. To adhere to legal and ethical standards, the identities of individual organisations and their specific data contributions are not disclosed. Cohorts are generated via the TriNetX user interface by applying inclusion and exclusion criteria. They can be matched for confounding factors and compared for outcomes of interest across specified time periods (Palchuk et al., 2023). Like other studies utilising the TriNetX network, this retrospective study is exempt from obtaining informed consent, and institutional ethical approval was not required. The data analysed is secondary data, does not involve intervention or interaction with human subjects, and has been de-identified in line with the de-identification standard outlined in Section §164.514(b)(1) of the HIPAA Privacy Rule (http://trinetx.com). The RECORD reporting guidelines were followed (Supplementary Table 1).

### 2.2. Definition of cohorts

#### 2.2.1. Level of systemic inflammation surrounding adversity

For the investigation of adverse life events, individuals aged 13-80 years were included if they had a record of a clinical encounter for examination and observation following a transport accident (Z04.1), work accident (Z04.2), other accident (Z04.3), alleged rape (Z04.4) or alleged physical abuse (Z04.7) between 1 January 2011 and 31 July 2023 (two years before accessing the data in August 2025). These codes were selected as they generally indicate an examination triggered by a reported event, rather than background psychosocial enquiry, and offer closer temporal proximity to the event onset than some other ICD-10 codes related to past traumatic experiences, especially abuse.

Peripheral leukocyte (white blood cell) counts were selected as the primary biomarker of systemic inflammation owing to the availability of the data and established use in previous studies (Danese et al., 2007; Penz et al., 2018; Surtees et al., 2003). Two cohorts were created: i) a mild inflammation cohort, in which individuals had leukocytes counts between 11.1 and 15.0 10^3^/µL (and no values ≥15.1 10^3^/µL) within one week before or after the medical encounter; and ii) a low inflammation cohort, where individuals had leukocyte counts between 4.0 and 11.0 10^3^/µL (and no values ≥11.1 10^3^/µL), consistent with the normal reference range, within the same time window (Fig. 1A). Thresholds were selected to differentiate normal from mildly elevated leukocyte counts while excluding individuals with severe chronic inflammatory states and other conditions associated with persistently and markedly increased leukocyte counts.

**Fig. 1.**
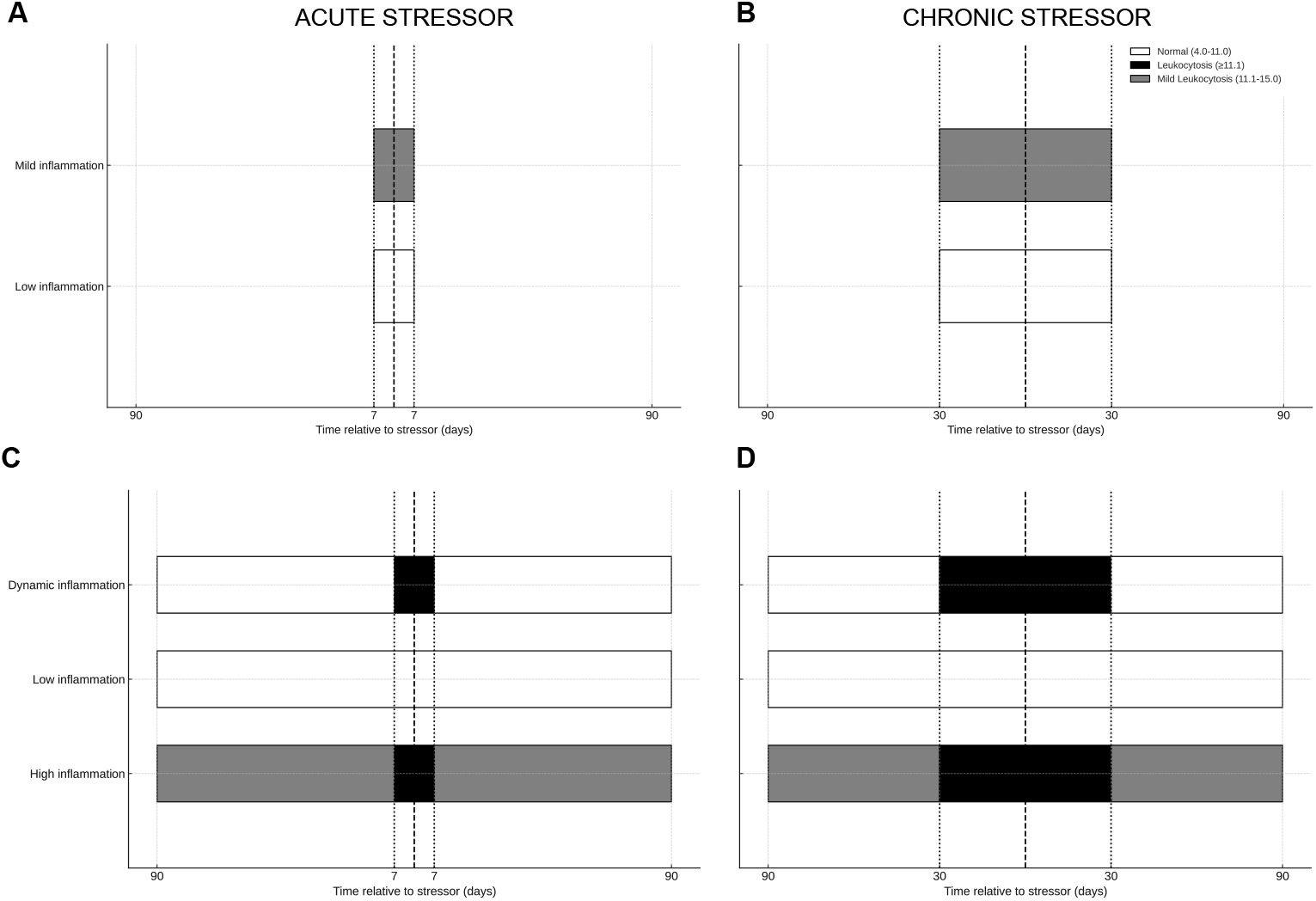
Time windows and leukocyte criteria for cohort definitions. For comparison of inflammation levels, cohorts were defined by either mild leukocytosis or normal leukocyte counts during the period within one week of an acute adverse event (A) or within one month of a record of a chronic stressor (B). For analyses of the dynamic inflammatory response versus persistent low or high inflammation, cohorts were defined by leukocyte counts within one week for acute stressors (C) or one month for chronic stressors (D), as well as counts before or after that period.

For the investigation of socioeconomic and psychosocial stressors, a contemporaneous cohort of individuals aged between 13 and 80 years was constructed with records of problems related to employment and unemployment (Z56), housing and economic circumstances (Z59), social environment (Z60), and other problems related to primary support group, including family circumstances (Z63). Because these stressors are more chronic and their recorded date is less likely to closely reflect their actual onset with precision, we applied a wider time window for capturing blood leukocyte data. Cases were included in the mild and low inflammation cohorts if they met the same criteria regarding leukocyte counts as above, but with the time window defined as one month before or after the first record of a stressor (Fig. 1B).

#### 2.2.2. Changes in systemic inflammation surrounding adversity

Cases were included if they had a record of examination following an adverse event or of a psychosocial stressor defined above. For the investigation of the systemic inflammatory profile surrounding clinical encounters following adverse life events, three cohorts were created: i) a dynamic inflammation cohort comprised of individuals with leukocytosis (≥11.1 10^3^/µL) within one week before or after the encounter, together with a normal leukocyte range (4.0-11.0 10^3^/µL) either between three months and one week before or between one week and three months after the encounter; ii) a low inflammation cohort including individuals with only normal leukocyte values in both the one-week window surrounding the encounter and the period up to three months before and after, with no higher values recorded; and iii) a high inflammation cohort including individuals with leukocytosis recorded within one week of the encounter, and mild leukocytosis (11.1-15.0 10^3^/µL) in the period up to three months before or after (Fig. 1C). In this analysis, no upper leukocyte threshold was applied within the immediate peri-adversity window to capture acute inflammatory responses while still excluding cases with severe persistent leukocytosis. Similar criteria were applied for psychosocial stressors, with time windows for leukocyte counts set at one month and up to three months before or after the recorded stressor (Fig. 1D).

### 2.3. Outcomes of interest

We compared pairs of cohorts for predefined outcomes of interest. These outcomes comprised records of diagnoses, selected a priori to represent a balanced overview across mental health conditions and interface disorders, linked to stress sensitivity and functional alterations, and their somatic counterparts. First, we included mental health and interface disorders, including anxiety, depression, PTSD or adjustment disorders, FND, somatoform disorders, sleep disorders, and cognitive symptoms. Second, we included somatic symptom diagnoses that represent pain, fatigue, sensory dysregulation, or autonomic disturbance, which often co-occur with the above disorders and are hypothesised to have overlapping biopsychosocial mechanisms and/or risk factors. This group comprised headache, chest pain, abdominal pain, low back pain, fibromyalgia, breathing abnormalities, palpitations, irritable bowel syndrome, pruritus, and malaise and fatigue. Outcomes of interest and the ICD-10 codes used are detailed in Supplementary Methods.

The time window for the analysis of outcomes extended between one week and two years after the record of an acute stressor, and from one month to two years after the record of a chronic stressor. For each outcome, individuals were excluded if they had a prior record of that outcome before the start of the analysis window.

### 2.4. Statistical analysis

Using measures of association, we calculated the odds ratios (ORs) for each outcome of interest comparing i) cohorts with mild inflammation to those with low levels of inflammation; and ii) cohorts with evidence of changes in systemic inflammation in the period surrounding adversity to those with persistent high and low inflammation. For each cohort comparison, *p* values for the 17 variables of interest were adjusted using the false discovery rate (FDR) method with *q* = 0.05.

For each analysis, cohort pairs were matched for demographic and clinical variables using propensity score matching. The algorithm used is integrated into the TriNetX platform and employs logistic regression to calculate propensity scores based on user-specified covariates. It then applies a greedy nearest-neighbour matching algorithm with a 1:1 ratio to pair cases and controls. We matched the cohorts for demographic variables, immune and haematological disorders, as well as infectious diseases and hospital or critical care treatment within the month before the record of the stressor (Supplementary Methods). Cohort pairs were additionally matched on variables defining the acute or chronic stressors. Although they were not actively matched for these variables, all cohort pairs were well-matched (SMD < 0.10) for records of common general medical conditions, common mental health disorders and groups of neurological conditions, as well as smoking status (Supplementary Tables 2-7).

We conducted additional sensitivity analyses. First, we examined the associations between leukocyte counts (and their changes around records of adversity) using a set of negative-control outcomes (lipoma, hallux valgus, ingrowing nail). In addition, we reassessed the association with outcomes of interest after excluding individuals with records of systemic glucocorticoid use during the observation period surrounding stressors (Supplementary Methods).

## 3. Results

### 3.1. Presence of systemic inflammation at the time of adversity

After matching, 36,772 individuals with records of examination following adverse events and mild leukocytosis and 36,772 with normal leukocyte counts were included in each cohort. A new diagnosis of an anxiety disorder was less common among cases with leukocytosis, as were cognitive symptoms, palpitations, fatigue, breathing abnormalities, and several pain types (Table 1, Fig. 2A).

**Table 1.** Incidence of a new outcome within one week to two years after the record of an adverse event in matched cohorts with mild leukocytosis and normal leukocyte counts. Numbers of cases (%) with an outcome are provided, along with the odds ratio (OR) and *p* values. *p* values significant after FDR correction for 17 variables are highlighted in bold.

| Outcome | N (%) with mild leukocytosis | N (%) with normal leukocytes | OR (95% CI) | *p* | *p* (FDR) |
| --- | --- | --- | --- | --- | --- |
| *Mental health/interface conditions* |  |  |  |  |  |
| Anxiety disorders | 1906 (6.4%) | 1993 (7.0%) | 0.914 (0.856-0.975) | 0.007 | **0.017** |
| Depression/depressive episode | 1761 (6.0%) | 1801 (6.4%) | 0.936 (0.875-1.002) | 0.058 | 0.090 |
| PTSD | 613 (1.7%) | 633 (1.8%) | 0.961 (0.859-1.075) | 0.482 | 0.482 |
| FND | 108 (0.3%) | 139 (0.4%) | 0.774 (0.602-0.996) | 0.046 | 0.078 |
| Somatoform disorders | 150 (0.4%) | 136 (0.4%) | 1.100 (0.872-1.388) | 0.423 | 0.449 |
| Sleep disorders | 391 (1.1%) | 436 (1.2%) | 0.891 (0.777-1.022) | 0.099 | 0.129 |
| Cognitive symptoms | 1561 (5.0%) | 1754 (5.7%) | 0.870 (0.811-0.933) | <0.001 | **0.002** |
| *Pain, somatic symptoms, fatigue* |  |  |  |  |  |
| Headache | 1562 (5.3%) | 1736 (6.3%) | 0.841 (0.784-0.902) | <0.001 | **0.002** |
| Unspecified chest pain | 1507 (5.4%) | 1635 (6.1%) | 0.878 (0.817-0.943) | <0.001 | **0.002** |
| Unspecified abdominal pain | 1442 (5.0%) | 1606 (5.7%) | 0.859 (0.798-0.924) | <0.001 | **0.002** |
| Low back pain | 1540 (5.2%) | 1613 (5.7%) | 0.907 (0.844-0.974) | 0.008 | **0.017** |
| Fibromyalgia | 181 (0.5%) | 199 (0.6%) | 0.904 (0.739-1.106) | 0.326 | 0.369 |
| Breathing abnormalities | 2032 (7.2%) | 2123 (7.7%) | 0.931 (0.874-0.992) | 0.026 | **0.049** |
| Palpitations | 499 (1.4%) | 629 (1.8%) | 0.782 (0.694-0.880) | <0.001 | **0.002** |
| Irritable bowel syndrome | 154 (0.4%) | 177 (0.5%) | 0.865 (0.697-1.074) | 0.188 | 0.228 |
| Pruritus | 343 (1.0%) | 387 (1.1%) | 0.880 (0.760-1.018) | 0.085 | 0.120 |
| Malaise and fatigue | 2065 (6.9%) | 2146 (7.5%) | 0.912 (0.857-0.971) | 0.004 | **0.011** |

**Fig. 2.**
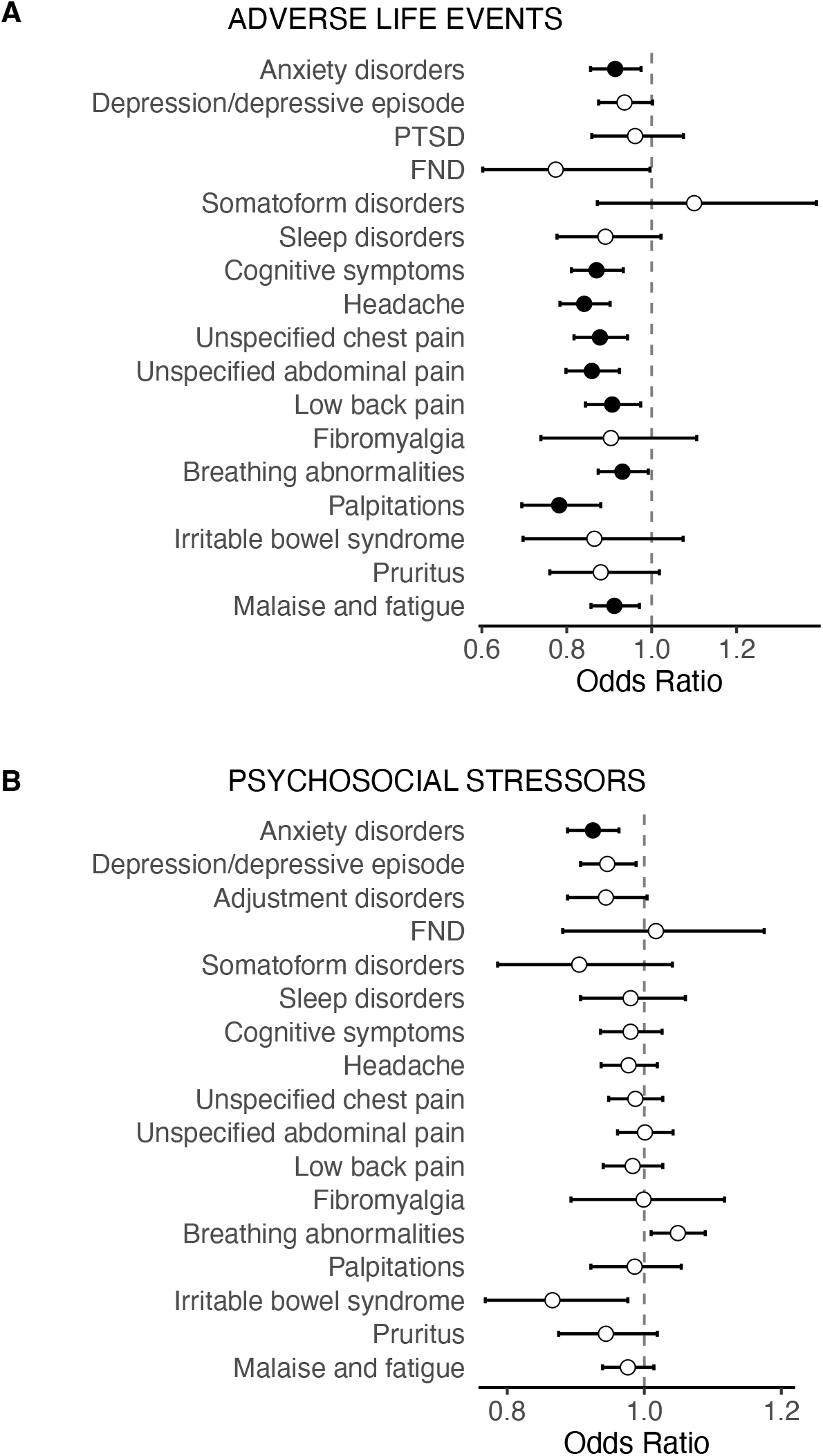
Odds ratios with 95% confidence intervals for new outcomes in individuals with mild versus low inflammation within two years following a recorded adverse life event (A) or psychosocial stressor (B). Filled circles indicate associations that remained significant after false discovery rate (FDR) correction. PTSD: post-traumatic stress disorder; FND: functional neurological disorder

Matched cohorts of individuals with mild leukocytosis around the first recorded psychosocial or socioeconomic stressor and those with normal leukocyte counts during the same period each included 87,936 cases. Following the record of the stressor, anxiety disorders were less common in cases with leukocytosis (Table 2, Fig. 2B).

**Table 2.** Incidence of a new outcome within one month to two years after the record of a psychosocial or socioeconomic stressor in matched cohorts with mild leukocytosis and normal leukocyte counts. Numbers of cases (%) with an outcome are provided, along with the odds ratio (OR) and *p* values. *p* values significant after FDR correction for 17 variables are highlighted in bold.

| Outcome | N (%) with mild leukocytosis | N (%) with normal leukocytes | OR (95% CI) | *p* | *p* (FDR) |
| --- | --- | --- | --- | --- | --- |
| *Mental health/interface conditions* |  |  |  |  |  |
| Anxiety disorders | 5166 (9.7%) | 5378 (10.4%) | 0.925 (0.888-0.963) | <0.001 | **0.009** |
| Depression/depressive episode | 4664 (9.3%) | 4681 (9.8%) | 0.946 (0.907-0.988) | 0.011 | 0.076 |
| Adjustment disorders | 2036 (2.6%) | 2150 (2.7%) | 0.944 (0.888-1.004) | 0.065 | 0.221 |
| FND | 377 (0.4%) | 370 (0.4%) | 1.017 (0.881-1.175) | 0.815 | 0.924 |
| Somatoform disorders | 373 (0.4%) | 412 (0.5%) | 0.905 (0.786-1.041) | 0.162 | 0.393 |
| Sleep disorders | 1270 (1.5%) | 1291 (1.5%) | 0.980 (0.907-1.060) | 0.618 | 0.808 |
| Cognitive symptoms | 3798 (5.0%) | 3890 (5.1%) | 0.980 (0.936-1.026) | 0.390 | 0.663 |
| *Pain, somatic symptoms, fatigue* |  |  |  |  |  |
| Headache | 4718 (6.7%) | 4774 (6.8%) | 0.977 (0.937-1.019) | 0.273 | 0.516 |
| Unspecified chest pain | 5129 (7.9%) | 5226 (8.0%) | 0.987 (0.948-1.027) | 0.512 | 0.725 |
| Unspecified abdominal pain | 4928 (7.3%) | 4990 (7.3%) | 1.001 (0.961-1.042) | 0.973 | 0.980 |
| Low back pain | 4194 (5.9%) | 4272 (6.0%) | 0.983 (0.940-1.027) | 0.433 | 0.669 |
| Fibromyalgia | 620 (0.7%) | 620 (0.7%) | 0.999 (0.893-1.117) | 0.980 | 0.980 |
| Breathing abnormalities | 5930 (9.7%) | 5853 (9.3%) | 1.049 (1.010-1.089) | 0.014 | 0.076 |
| Palpitations | 1751 (2.1%) | 1762 (2.1%) | 0.986 (0.922-1.054) | 0.676 | 0.821 |
| Irritable bowel syndrome | 505 (0.6%) | 582 (0.7%) | 0.866 (0.768-0.976) | 0.018 | 0.076 |
| Pruritus | 1308 (1.5%) | 1381 (1.6%) | 0.944 (0.875-1.019) | 0.137 | 0.388 |
| Malaise and fatigue | 5745 (8.7%) | 5803 (8.9%) | 0.976 (0.939-1.014) | 0.206 | 0.438 |

### 3.2. Change in inflammatory state in the period surrounding adversity

We compared matched cohorts of 12,303 cases with dynamic changes in leukocyte counts to those with normal leukocytes, and matched cohorts of 12,874 cases with dynamic changes in leukocyte counts to those with persistent leukocytosis. Compared to those with persistent low systemic inflammation, cases with dynamic inflammation in the period surrounding the adverse event had decreased rates of subsequent anxiety, depression, FND, cognitive symptoms, sleep disorders, fatigue, various somatic symptoms, and conditions associated with pain. Compared to the cohort with high inflammation, those with dynamic changes had less subsequent anxiety, depression, PTSD, cognitive symptoms, sleep disorders, several somatic symptoms, fatigue and pain (Table 3, Fig. 3A).

**Table 3.** Incidence of a new outcome within one week to two years after the record of an adverse event in matched cohorts with leukocytosis only surrounding adversity versus persistent normal or high leukocyte counts. Numbers of cases (%) with an outcome are provided, along with the odds ratio (OR) and *p* values. *p* values significant after FDR correction (17 variables per comparison) are highlighted in bold.

| Outcome | DYNAMIC vs. LOW INFLAMMATION<br>(N = 12303) |  |  |  |  | DYNAMIC vs. HIGH INFLAMMATION<br>(N = 12874) |  |  |  |  |
| --- | --- | --- | --- | --- | --- | --- | --- | --- | --- | --- |
|  | N (%) with leukocyte count change | N (%) with normal leukocytes | OR (95% CI) | *p* | *p* (FDR) | N (%) with leukocyte count change | N (%) with high leukocytes | OR (95% CI) | *p* | *p* (FDR) |
| *Mental health/interface conditions* |  |  |  |  |  |  |  |  |  |  |
| Anxiety disorders | 976 (11.4%) | 1038 (13.3%) | 0.839 (0.764-0.921) | <0.001 | **0.001** | 1028 (11.4%) | 1257 (13.7%) | 0.810 (0.742-0.885) | <0.001 | **0.001** |
| Depression/depressive episode | 915 (11.0%) | 993 (13.3%) | 0.806 (0.732-0.887) | <0.001 | **0.001** | 944 (10.8%) | 1197 (13.4%) | 0.783 (0.715-0.857) | <0.001 | **0.001** |
| PTSD | 287 (2.5%) | 291 (2.5%) | 0.975 (0.826-1.150) | 0.762 | 0.762 | 313 (2.6%) | 375 (3.1%) | 0.831 (0.714-0.968) | 0.017 | **0.032** |
| FND | 56 (0.5%) | 110 (0.9%) | 0.503 (0.364-0.695) | <0.001 | **0.001** | 63 (0.5%) | 77 (0.6%) | 0.817 (0.585-1.141) | 0.235 | 0.307 |
| Somatoform disorders | 103 (0.9%) | 81 (0.7%) | 1.266 (0.945-1.695) | 0.113 | 0.128 | 109 (0.9%) | 132 (1.0%) | 0.825 (0.639-1.064) | 0.137 | 0.212 |
| Sleep disorders | 231 (2.0%) | 286 (2.5%) | 0.799 (0.671-0.952) | 0.012 | **0.017** | 241 (1.9%) | 296 (2.4%) | 0.815 (0.687-0.968) | 0.020 | **0.034** |
| Cognitive symptoms | 943 (10.2%) | 1185 (13.3%) | 0.742 (0.677-0.812) | <0.001 | **0.001** | 960 (10.0%) | 1409 (15.1%) | 0.624 (0.572-0.681) | <0.001 | **0.001** |
| *Pain, somatic symptoms, fatigue* |  |  |  |  |  |  |  |  |  |  |
| Headache | 817 (9.0%) | 1031 (12.6%) | 0.688 (0.624-0.758) | <0.001 | **0.001** | 849 (8.9%) | 914 (9.3%) | 0.956 (0.866-1.054) | 0.363 | 0.441 |
| Unspecified chest pain | 828 (10.4%) | 943 (13.5%) | 0.750 (0.679-0.828) | <0.001 | **0.001** | 869 (10.4%) | 993 (11.7%) | 0.873 (0.792-0.961) | 0.006 | **0.013** |
| Unspecified abdominal pain | 783 (9.1%) | 972 (12.3%) | 0.714 (0.646-0.789) | <0.001 | **0.001** | 814 (9.0%) | 1133 (12.4%) | 0.697 (0.634-0.767) | <0.001 | **0.001** |
| Low back pain | 794 (8.9%) | 876 (10.7%) | 0.817 (0.738-0.904) | <0.001 | **0.001** | 823 (8.7%) | 834 (8.7%) | 1.006 (0.910-1.113) | 0.904 | 0.904 |
| Fibromyalgia | 90 (0.8%) | 118 (1.0%) | 0.747 (0.567-0.984) | 0.037 | **0.045** | 90 (0.7%) | 108 (0.9%) | 0.833 (0.629-1.103) | 0.200 | 0.283 |
| Breathing abnormalities | 1034 (14.1%) | 1228 (18.1%) | 0.741 (0.677-0.811) | <0.001 | **0.001** | 1075 (13.9%) | 1549 (20.8%) | 0.618 (0.568-0.673) | <0.001 | **0.001** |
| Palpitations | 253 (2.2%) | 367 (3.3%) | 0.662 (0.563-0.779) | <0.001 | **0.001** | 256 (2.1%) | 268 (2.2%) | 0.959 (0.806-1.140) | 0.633 | 0.673 |
| Irritable bowel syndrome | 90 (0.8%) | 107 (0.9%) | 0.832 (0.628-1.102) | 0.199 | 0.211 | 91 (0.7%) | 100 (0.8%) | 0.909 (0.683-1.208) | 0.510 | 0.578 |
| Pruritus | 188 (1.6%) | 233 (2.0%) | 0.789 (0.650-0.958) | 0.016 | **0.021** | 192 (1.6%) | 266 (2.2%) | 0.716 (0.594-0.863) | <0.001 | **0.001** |
| Malaise and fatigue | 1213 (14.8%) | 1321 (18.1%) | 0.787 (0.723-0.858) | <0.001 | **0.001** | 1286 (14.8%) | 1611 (18.2%) | 0.779 (0.719-0.844) | <0.001 | **0.001** |

**Fig. 3.**
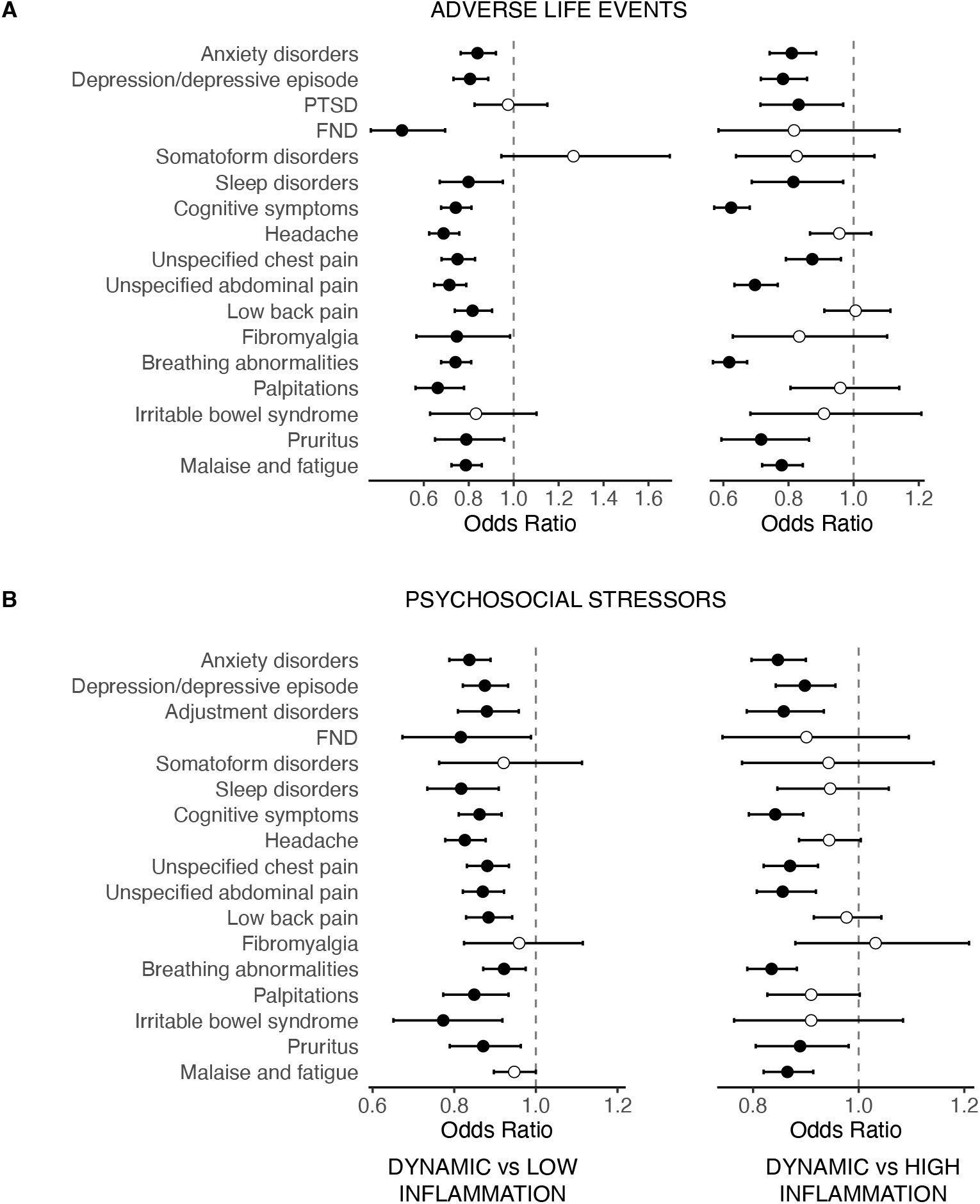
Odds ratios with 95% confidence intervals for new outcomes in individuals with dynamic versus low inflammation (left) and dynamic versus high inflammation (right) within two years following a recorded adverse life event (A) or psychosocial stressor (B). Filled circles indicate associations that remained significant after false discovery rate (FDR) correction. PTSD: post-traumatic stress disorder; FND: functional neurological disorder

For psychosocial stressors, we compared matched cohorts of 31,447 cases with dynamic changes to those with normal leukocyte counts, as well as matched cohorts of 30,842 cases with dynamic changes to those with persistent leukocytosis. Compared to the low- and high-inflammation cohorts, the cohort with a dynamic inflammatory response exhibited lower rates of anxiety, depression, adjustment disorders, cognitive symptoms, fatigue, and various pain-related and other somatic conditions. Sleep disorders and FND were less common in individuals with a dynamic inflammatory response compared to those with persistent low inflammation (Table 4, Fig. 3B).

**Table 4.** Incidence of a new outcome within one month to two years after the record of a psychosocial or socioeconomic stressor in matched cohorts with leukocytosis only surrounding adversity versus persistent normal or high leukocyte counts. Numbers of cases (%) with an outcome are provided, along with the odds ratio (OR) and *p* values. *p* values significant after FDR correction (17 variables per comparison) are highlighted in bold.

| Outcome | DYNAMIC vs. LOW INFLAMMATION<br>(N = 31447) |  |  |  |  | DYNAMIC vs. HIGH INFLAMMATION<br>(N = 30842) |  |  |  |  |
| --- | --- | --- | --- | --- | --- | --- | --- | --- | --- | --- |
|  | N (%) with leukocyte count change | N (%) with normal leukocytes | OR (95% CI) | *p* | *p* (FDR) | N (%) with leukocyte count change | N (%) with high leukocytes | OR (95% CI) | *p* | *p* (FDR) |
| *Mental health/interface conditions* |  |  |  |  |  |  |  |  |  |  |
| Anxiety disorders | 2376 (13.6%) | 2621 (15.8%) | 0.837 (0.788-0.889) | <0.001 | **0.001** | 2321 (13.7%) | 2593 (15.8%) | 0.847 (0.797-0.900) | <0.001 | **0.001** |
| Depression/depressive episode | 2184 (13.1%) | 2248 (14.7%) | 0.875 (0.821-0.932) | <0.001 | **0.001** | 2170 (13.3%) | 2374 (14.6%) | 0.898 (0.843-0.956) | <0.001 | **0.001** |
| Adjustment disorders | 1066 (3.8%) | 1197 (4.4%) | 0.880 (0.809-0.958) | 0.003 | **0.005** | 1031 (3.8%) | 1188 (4.4%) | 0.858 (0.788-0.934) | <0.001 | **0.001** |
| FND | 192 (0.6%) | 234 (0.8%) | 0.816 (0.673-0.988) | 0.037 | **0.045** | 194 (0.6%) | 215 (0.7%) | 0.901 (0.742-1.095) | 0.296 | 0.387 |
| Somatoform disorders | 209 (0.7%) | 226 (0.7%) | 0.921 (0.763-1.113) | 0.395 | 0.420 | 205 (0.7%) | 217 (0.7%) | 0.943 (0.779-1.142) | 0.549 | 0.583 |
| Sleep disorders | 624 (2.1%) | 753 (2.5%) | 0.817 (0.734-0.909) | <0.001 | **0.001** | 615 (2.1%) | 648 (2.2%) | 0.946 (0.846-1.057) | 0.328 | 0.398 |
| Cognitive symptoms | 2106 (8.3%) | 2391 (9.5%) | 0.862 (0.811-0.916) | <0.001 | **0.001** | 2091 (8.5%) | 2428 (10.0%) | 0.842 (0.792-0.895) | <0.001 | **0.001** |
| *Pain, somatic symptoms, fatigue* |  |  |  |  |  |  |  |  |  |  |
| Headache | 2221 (9.5%) | 2542 (11.3%) | 0.826 (0.778-0.877) | <0.001 | **0.001** | 2164 (9.5%) | 2293 (10.0%) | 0.944 (0.887-1.004) | 0.065 | 0.100 |
| Unspecified chest pain | 2459 (12.2%) | 2659 (13.6%) | 0.881 (0.831-0.934) | <0.001 | **0.001** | 2391 (12.3%) | 2675 (13.9%) | 0.870 (0.820-0.923) | <0.001 | **0.001** |
| Unspecified abdominal pain | 2470 (11.7%) | 2750 (13.2%) | 0.870 (0.821-0.922) | <0.001 | **0.001** | 2412 (11.7%) | 2689 (13.4%) | 0.856 (0.807-0.919) | <0.001 | **0.001** |
| Low back pain | 2004 (8.6%) | 2190 (9.6%) | 0.884 (0.829-0.942) | <0.001 | **0.001** | 1934 (8.4%) | 1964 (8.6%) | 0.977 (0.915-1.043) | 0.482 | 0.546 |
| Fibromyalgia | 334 (1.1%) | 344 (1.2%) | 0.959 (0.824-1.115) | 0.585 | 0.585 | 314 (1.1%) | 304 (1.0%) | 1.032 (0.880-1.209) | 0.700 | 0.700 |
| Breathing abnormalities | 2794 (15.6%) | 2983 (16.7%) | 0.922 (0.871-0.975) | 0.005 | **0.007** | 2766 (16.2%) | 3039 (18.8%) | 0.835 (0.789-0.883) | <0.001 | **0.001** |
| Palpitations | 838 (2.9%) | 966 (3.4%) | 0.849 (0.773-0.933) | 0.001 | **0.002** | 818 (2.9%) | 899 (3.2%) | 0.910 (0.827-1.002) | 0.055 | 0.094 |
| Irritable bowel syndrome | 234 (0.8%) | 300 (1.0%) | 0.773 (0.651-0.918) | 0.003 | **0.005** | 242 (0.8%) | 266 (0.9%) | 0.910 (0.764-1.084) | 0.293 | 0.387 |
| Pruritus | 745 (2.5%) | 844 (2.9%) | 0.871 (0.789-0.963) | 0.007 | **0.009** | 770 (2.6%) | 858 (3.0%) | 0.889 (0.805-0.981) | 0.019 | **0.036** |
| Malaise and fatigue | 3038 (15.2%) | 3072 (15.9%) | 0.947 (0.897-1.000) | 0.051 | 0.058 | 3005 (15.7%) | 3255 (17.7%) | 0.865 (0.820-0.914) | <0.001 | **0.001** |

### 3.3. Sensitivity analyses

Measures of static or dynamic inflammatory response in the period surrounding stressor records were not associated with negative-control outcomes (Supplementary Tables 8 and 9). After excluding cases with systemic glucocorticoid treatment, some qualitative and quantitative differences emerged. However, the overall pattern remained unchanged. Mild leukocytosis in the period surrounding adverse life events or psychosocial stressors continued to be associated with a more favourable risk profile compared with normal leukocyte counts, and dynamic changes in leukocyte counts similarly remained linked to a reduced risk of subsequent psychiatric or somatic outcomes (Supplementary Tables 10-13).

## 4. Discussion

Our large EHR-based retrospective cohort study provides evidence that the pattern of systemic inflammatory response surrounding adverse life events is associated with long-term risk of mental illness and somatic symptom diagnoses over the subsequent two years, in line with previous evidence. However, individuals with mild leukocytosis in the period surrounding stressor exposure, and particularly those showing dynamic changes in leukocyte counts, had lower risk of several outcomes, including anxiety, depression, sleep disturbance, cognitive symptoms, fatigue, pain syndromes, somatic symptoms, and FND. These findings suggest that a dynamic inflammatory response marks greater homeostatic capacity or physiological adaptability, and is associated with fewer sequelae of adversity.

Anxiety disorders showed the most consistent risk reduction in individuals with mild inflammation (compared to those with no evidence of systemic inflammation), and among individuals with dynamic inflammatory changes (compared to those with persistently low and persistently elevated inflammatory profiles), both following adverse life events and psychosocial stressors. Depression and adjustment disorders were also less common in those with dynamic immune responses, suggesting a broader protective effect. Interestingly, PTSD was less common in individuals with dynamic inflammatory response compared to those with persistent high inflammation, but not compared to those with low inflammation, contrasting with previous findings of reduced TNF-α and IFN-γ in individuals at greater risk for PTSD (Michopoulos et al., 2020). This discrepancy may reflect the relative rarity of PTSD, limiting statistical power, or distinct biological mechanisms (potentially including inflammatory components) that are not captured by peripheral leukocyte counts. The absence of consistent associations for PTSD suggests that although dynamic systemic inflammation may protect against a broad array of neuropsychiatric conditions, it is unlikely to be the sole determinant of resilience.

Inflammatory illnesses may be associated with the onset of FND, as the accompanying distress, physiological arousal, inflammatory and interoceptive changes may reinforce maladaptive symptom perception mechanisms. However, the specific role of inflammatory profiles in FND remains poorly understood (Paredes-Echeverri et al., 2022). Following both adverse life events and psychosocial stressors, we observed a lower risk of FND in individuals with a dynamic inflammatory response compared to those showing persistently low inflammation. FND can be understood as a disorder of how the brain predicts and interprets bodily and environmental signals (Edwards et al., 2012). A transient and well-regulated inflammatory response may act as a salient interoceptive signal that promotes updating these predictions after stress, thereby recalibrating stress-related circuits and supporting recovery. When this immune response is blunted, inaccurate expectations about threat and bodily symptoms may persist, increasing vulnerability to FND and related mental health outcomes. Importantly, such blunting may not simply reflect reduced inflammatory signalling, but a broader predisposition to ineffective processing of stressors. The immune system balances two complementary strategies: resistance, which aims to eliminate a stressor, and tolerance, which aims to limit the harm caused by the response to the organism (Medzhitov et al., 2012). A blunted inflammatory response may reflect failure of both arms: insufficient resistance to mount a robust acute response, or inadequate tolerance to adaptively regulate the system afterwards. Conversely, when inflammation is excessive or prolonged, prediction systems may be biased towards hypervigilance, perpetuating chronic symptoms.

Several outcomes, including sleep disorders, cognitive symptoms, fatigue, and pain syndromes, were strongly influenced by the nature of the inflammatory response, with dynamic changes conferring reduced risk compared with both persistently low and persistently elevated inflammatory profiles. The observed pattern suggests that an adaptive immune response helps maintain homeostatic regulation across the mind-body systems. This pattern, however, was not consistent across all investigated outcomes. While individual diagnoses of somatic symptoms, including pain, fatigue, palpitations, breathing abnormalities, and pruritus, were less frequent in the dynamic group, no corresponding reduction in diagnosed somatoform disorders was observed. As noted for PTSD, this may reflect the relative rarity of this diagnosis, but could also be due to the limited follow-up period after the stressor, which may not have been sufficient for the diagnosis to be recognised.

Several prospective studies have reported that an elevated inflammatory response following a traumatic experience is associated with a greater risk of subsequent mental health disorders (Pervanidou et al., 2007; Robles et al., 2024; von Känel et al., 2022). At first glance, these findings may appear at odds with the results of the current study, in which longitudinal inflammatory profiles were more informative than absolute levels at a single time point. However, single-time-point assessments may fail to distinguish transient, context-appropriate inflammatory responses from later dysregulated or blunted immune states. Evidence that insufficient early inflammatory signalling predicts transition from acute to chronic pain (Parisien et al., 2022) further suggests the importance of immune dynamics rather than absolute levels. Taken together, these findings highlight the importance of timing, context, and regulatory dynamics: both a pre-existing elevated inflammatory state and a blunted acute immune response to trauma may confer risk (Katrinli et al., 2022). Our results support the view that mental health trajectories depend on precisely calibrated post-stress immune responses, with risk arising when the balance is disrupted in either direction.

Given the bidirectional nature of brain-immune communication (Dantzer et al., 2018), psychological resilience, effective coping strategies, and social support may enhance the habituation of the immune response following acute stress (Coan et al., 2006; Cohen et al., 2015). A dynamic inflammatory response may therefore reflect not only intrinsic immune properties, but also higher-order regulatory processes that shape stress recovery. Building on this framework, several interconnected mechanisms may underlie the protective effect of an acute, self-resolving inflammatory response against later stress-related outcomes. One is the concept of behavioural immunisation, which posits that transient activation of immune and stress systems during an acute challenge facilitates adaptive recalibration of neuroimmune networks, enhancing resilience to subsequent stressors (Lewitus & Schwartz, 2009). In addition, individuals who exhibit a robust but self-limiting activation of the hypothalamic-pituitary-adrenal axis may develop attenuated inflammatory responses to later stressors, consistent with adaptive calibration of stress-regulatory systems through allostatic processes. Such adaptation may reduce the likelihood of reactivation when encountering repeated or familiar stressors (Thoma et al., 2017). A third mechanism involves the concept of tolerance, a complementary process to allostasis, which highlights the system’s ability to minimise induced damage and restore functional homeostasis (Medzhitov et al., 2012).

Strengths of our study include the use of very large, well-matched real-world geographically diverse cohorts, which enabled the examination of a broad range of mental health and somatic symptom outcomes with substantial statistical power. The longitudinal structure of the data allowed us to move beyond single-timepoint measures of inflammation and instead examine inflammatory trajectories in relation to subsequent outcomes. The replication of associations across both adverse life events and psychosocial stressors further strengthens the robustness and generalisability of our findings. Nevertheless, several limitations must be considered. First, leukocyte counts provide a relatively crude measure of inflammation, lacking specificity regarding the underlying immune pathways involved. Future research should aim to incorporate more detailed immunological measures, encompassing other systemic markers, specific cytokine profiles, and markers of immune cell activation. These were either unavailable or too infrequent in the TriNetX dataset to be used in this study. Second, residual confounding cannot be excluded, particularly from the nature of healthcare encounters, as well as unmeasured psychosocial factors that may influence both systemic inflammation and mental health outcomes. Third, our assessment of the dynamic nature of the inflammatory response relied on two leukocyte measurements, which failed to capture the full trajectory of immune activation and resolution. It is therefore possible that inflammation remained unresolved in some cases. However, adding additional data points would substantially restrict the number of cases included. Further, the temporal resolution in the EHR is limited, particularly for establishing the timing of exposure to psychosocial stressors. It is also difficult to disentangle whether the inflammation was pre-existing or triggered by the event itself. In addition, we applied a uniform statistical approach across disorders that vary in their temporal relationship to adversity and their long-term trajectories, which may have limited our ability to observe associations for certain disorders. Further, TriNetX primarily captures data from U.S. centres and from insured individuals treated mostly in academic and acute care settings, which may not fully represent broader demographic groups (Nassar et al., 2025). Finally, as with all observational studies, causal inferences cannot be drawn, and interventional studies targeting inflammatory dynamics are required to confirm a potential protective effect of a dynamic inflammatory response.

Our findings challenge the prevailing view that inflammation surrounding stressors is a predominantly harmful mediator of pathology. They align with emerging evidence suggesting that a well-regulated inflammatory response to adverse life events or psychosocial stressors – or the greater homeostatic capacity it might reflect – may facilitate adaptive recovery processes, thereby contributing to more favourable long-term health outcomes. Our results highlight the need to refine our understanding of how the immune system responds to stress. Rather than viewing inflammation as a static risk factor, future research should prioritise longitudinal study designs capable of a more nuanced appreciation of timing, context, and individual variability of the immune response (Moriarity & Slavich, 2023). Future studies should also consider the notion that measurement of inflammatory markers may be telling us more about the status of an individual’s immune system than providing a direct readout of a mechanistically pathogenic disease process. In turn, immune system dynamics may have far broader implications for whole-body inflammation, tissue regulation, and homeostasis than current approaches allow for. Understanding how these dynamic processes shape brain-body interactions in the context of adversity may clarify why similar stressors yield divergent outcomes between individuals and could point toward novel intervention targets.

## Data Availability

All data produced in the present study are available upon reasonable request to the authors.

## Acknowledgements

The authors thank TriNetX for providing access to the platform, without constraints on the analyses conducted or the decision to publish

