## Supplementary Material for "Dynamics of the systemic inflammatory response surrounding life events and the association with neuropsychiatric and somatic outcomes"

This appendix forms part of the submission of the manuscript:

### SUPPLEMENTARY METHODS

#### 1. Definition of outcomes of interest

The following variables of interest were identified among mental health/interface conditions, each encoded as a dichotomous variable:
1) **Anxiety disorders,** encoded as Other anxiety disorders (F41)
2) **Depression/depressive episode**, encoded as Depressive episode (F32) or Major depressive disorder, recurrent (F33)
3) **Post-traumatic stress disorder (PTSD)** (F43.1) for acute stressors; or **Adjustment disorders** (F43.2) for chronic stressors
4) **Functional neurological disorder (FND)**, encoded as Conversion disorder with motor symptom of deficit (F44.4), Conversion disorder with seizures or convulsions (F44.5), Conversion disorder with sensory symptom or deficit (F44.6), Conversion disorder with mixed symptom presentation (F44.7), Other dissociative and conversion disorders (F44.89), or Dissociative and conversion disorder, unspecified (F44.9)
5) **Somatoform disorders** (F45)
6) **Sleep disorders,** encoded as Sleep disorders not due to a substance or known physiological condition (F51)
7) **Cognitive symptoms,** encoded as Other symptoms and signs involving cognitive functions and awareness (R41.8)

Among conditions and symptoms associated with pain, somatic symptoms and fatigue, the following outcomes were of interest (each encoded as a dichotomous variable):
8) **Headache** (R51)
9) **Unspecified chest pain,** encoded as Chest pain, unspecified (R07.9)
10) **Unspecified abdominal pain** (R10.9)
11) **Low back pain** (M54.5)
12) **Fibromyalgia** (M79.7)
13) **Breathing abnormalities,** encoded as Abnormalities of breathing (R06)
14) **Palpitations** (R00.2)
15) **Irritable bowel syndrome** (K58)
16) **Pruritus**, encoded as Pruritus, unspecified (L29.9)
17) **Malaise and fatigue** (R53)

#### 2. Definition of covariates used for matching

Propensity score matching was performed on 26 characteristics, including demographic variables, comorbidities affecting immune response and leukocyte counts, and hospitalisation within the month between the record of a stressor:
1) **Age** at index event (record of a stressor)
2) **Gender,** encoded as three separate dichotomous variables: Female, Male, or Unknown
3) **Race,** encoded as seven separate dichotomous variables: White, Black or African American, Asian, American Indian or Alaska Native, Native Hawaiian or Other Pacific Islander, Other Race, Unknown Race
4) **Ethnicity,** encoded as Hispanic or Latino, not Hispanic or Latino, Unknown Ethnicity
5) **Diseases of the blood and blood-forming organs and certain disorders involving the immune mechanism** as a dichotomous variable (D50-D89) and additionally as a sub-category Certain disorders involving the immune mechanism (D80-D89)
6) **Certain infectious and parasitic diseases** (A00-B99)

7) **Systemic connective tissue disorders** (M30-M36)
8) **Inflammatory polyarthropathies** (M05-M14)
9) **Malignant neoplasms of lymphoid, hematopoietic and related tissue** (C81-C96)
10) **Hospital inpatient and observation care services**
11) **Critical care services**
12) Groups were matched for stressor types. For the investigation of psychosocial stressors, these were: **Problems related to social environment, Other problems related to primary support group, including family circumstances, Problems related to housing and economic circumstances, Problems related to employment and unemployment characteristic(s)**. For the investigation of acute adverse events, these were: **Encounter for examination and observation following transport accident, Encounter for examination and observation following work accident, Encounter for examination and observation following other accident, Encounter for examination and observation following alleged rape, Encounter for examination and observation following alleged physical abuse.**

### 3. Sensitivity analyses

We conducted sensitivity analyses to verify the association between the inflammatory response and long-term outcomes: 1) we verified the association using negative control outcomes, and 2) we excluded individuals with records of glucocorticoid therapy.

#### *3.1. Negative control outcomes*

To test the specificity of our findings, we repeated the analyses described above using a set of outcomes for which no meaningful association with an inflammatory response would be expected**:**
1) **Hallux valgus**, encoded as Hallux valgus, acquired (M20.1)

2) **Lipoma,** encoded as Benign lipomatous neoplasm (D17)
3) **Ingrown nail** (L60.0)

These conditions were selected as negative controls as they are common, largely non-inflammatory, and not known to fluctuate in relation to adverse life events, psychosocial stressors or the activation of the immune system.

#### *3.2 Excluding cases with glucocorticoid therapy*

As glucocorticoid treatment may influence both the systemic inflammatory response and leukocyte counts, we repeated the analyses described above after excluding individuals with evidence of systemic corticosteroid use. TriNetX provides only limited information on treatment context, dosage or duration that would not be possible to interrogate within our main study design. In addition, EHR-based medication lists often contain outdated or discontinued medications, which may lead to past medications being recorded as current. However, to address the uncertainty of the role of corticosteroids, we opted for a deliberately stringent approach in the sensitivity analysis. We excluded all individuals with any record of systemic glucocorticoid use within the period of leukocyte count observations, by excluding records with the Anatomical Therapeutic Classification (ATC) code H02AB. The same approach to matching was applied as in the primary analysis.

**Supplementary Table 1. The RECORD statement.** Checklist of items, extended from the STROBE statement, that should be reported in observational studies using routinely collected health data.

| | Item No. | STROBE items | Location in manuscript where items are reported | RECORD items | Location in manuscript where items are reported |
|---|---|---|---|---|---|
| **Title and abstract** | | | | | |
| | 1 | (a) Indicate the study's design with a commonly used term in the title or the abstract (b) Provide in the abstract an informative and balanced summary of what was done and what was found | Abstract | RECORD 1.1: The type of data used should be specified in the title or abstract. When possible, the name of the databases used should be included.<br><br>RECORD 1.2: If applicable, the geographic region and timeframe within which the study took place should be reported in the title or abstract.<br><br>RECORD 1.3: If linkage between databases was conducted for the study, this should be clearly stated in the title or abstract. | Abstract<br><br>N/A<br><br>N/A |
| **Introduction** | | | | | |
| Background rationale | 2 | Explain the scientific background and rationale for the investigation being reported | Abstract, Introduction | | |
| Objectives | 3 | State specific objectives, including any prespecified hypotheses | Introduction | | |
| **Methods** | | | | | |
| Study Design | 4 | Present key elements of study design early in the paper | Abstract, Introduction, Methods | | |
| Setting | 5 | Describe the setting, locations, and relevant dates, including periods of recruitment, exposure, follow-up, and data collection | Methods (2.1, 2.2, 2.3), Supplementary Methods | | |
| Participants | 6 | *(a) Cohort study* - Give the eligibility criteria, and the sources and methods of selection of participants. Describe methods of follow-up<br>*Case-control study* - Give the eligibility criteria, and the sources and methods of case ascertainment and control selection. Give the rationale for the choice of cases and controls<br>*Cross-sectional study* - Give the eligibility criteria, and the sources and methods of selection of participants<br><br>*(b) Cohort study* - For matched studies, give matching criteria and number of exposed and unexposed<br>*Case-control study* - For matched studies, give matching criteria and the number of controls per case | Methods (2.1, 2.2, 2.3), Fig. 1, Supplementary Methods<br><br>N/A | RECORD 6.1: The methods of study population selection (such as codes or algorithms used to identify subjects) should be listed in detail. If this is not possible, an explanation should be provided.<br><br>RECORD 6.2: Any validation studies of the codes or algorithms used to select the population should be referenced. If validation was conducted for this study and not published elsewhere, detailed methods and results should be provided.<br><br>RECORD 6.3: If the study involved linkage of databases, consider use of a flow diagram or other graphical display to demonstrate the data linkage process, including the number of individuals with linked data at each stage. | Methods (2.1, 2.2), Fig. 1, Supplementary Methods<br><br>Methods (2.1, 2.4), Supplementary Methods, Supplementary Tables 9-12<br><br>N/A |
| Variables | 7 | Clearly define all outcomes, exposures, predictors, potential confounders, and effect | Methods (2.2, 2.3, 2.4), Supplementary Methods | RECORD 7.1: A complete list of codes and algorithms used to classify exposures, outcomes, confounders, and | Methods (2.2, 2.3, 2.4), Supplementary Methods |

| | | | | | |
|---|---|---|---|---|---|
| | | modifiers. Give diagnostic criteria, if applicable. | | effect modifiers should be provided. If these cannot be reported, an explanation should be provided. | |
| Data sources/ measurement | 8 | For each variable of interest, give sources of data and details of methods of assessment (measurement).<br>Describe comparability of assessment methods if there is more than one group | Methods (2.2, 2.3), Supplementary Methods | | |
| Bias | 9 | Describe any efforts to address potential sources of bias | Methods (2.1, 2.4), Suplementary Methods | | |
| Study size | 10 | Explain how the study size was arrived at | Methods (2.2) | | |
| Quantitative variables | 11 | Explain how quantitative variables were handled in the analyses. If applicable, describe which groupings were chosen, and why | Methods (2.2, 2.3, 2.4), Supplementary Methods | | |
| Statistical methods | 12 | (a) Describe all statistical methods, including those used to control for confounding<br>(b) Describe any methods used to examine subgroups and interactions<br>(c) Explain how missing data were addressed<br>(d) *Cohort study* - If applicable, explain how loss to follow-up was addressed<br>*Case-control study* - If applicable, explain how matching of cases and controls was addressed<br>*Cross-sectional study* - If applicable, describe analytical methods taking account of sampling strategy<br>(e) Describe any sensitivity analyses | Methods (2.4), Supplementary Methods | | |
| Data access and cleaning methods | | .. | Methods (2.1), Supplementary Methods | RECORD 12.1: Authors should describe the extent to which the investigators had access to the database population used to create the study population.<br><br>RECORD 12.2: Authors should provide information on the data cleaning methods used in the study. | Methods (2.1), Acknowledgements<br><br>Methods (2.1), Supplementary Methods |
| Linkage | | .. | N/A | RECORD 12.3: State whether the study included person-level, institutional-level, or other data linkage across two or more databases. The methods of linkage and methods of linkage quality evaluation should be provided. | N/A |
| **Results** | | | | | |
| Participants | 13 | (a) Report the numbers of individuals at each stage of the study (*e.g.*, numbers potentially eligible, examined for eligibility, confirmed eligible, included in the study, completing follow-up, and analysed)<br>(b) Give reasons for non-participation at each stage.<br>(c) Consider use of a flow diagram | Results (3.1, 3.2, 3.3), Tables 1-4, Supplementary Tables 2-13 | RECORD 13.1: Describe in detail the selection of the persons included in the study (*i.e.,* study population selection) including filtering based on data quality, data availability and linkage. The selection of included persons can be described in the text and/or by means of the study flow diagram. | Methods, Tables 1-4, , Supplementary Tables 2-13 |
| Descriptive data | 14 | (a) Give characteristics of study participants (*e.g.*, demographic, clinical, social) and | Tables 1-4, Supplementary Tables 2-13 | | |

| | | | | | |
|---|---|---|---|---|---|
| | | information on exposures and potential confounders<br>(b) Indicate the number of participants with missing data for each variable of interest<br>(c) *Cohort study* - summarise follow-up time (*e.g.*, average and total amount) | | | |
| Outcome data | 15 | *Cohort study* - Report numbers of outcome events or summary measures over time<br>*Case-control study* - Report numbers in each exposure category, or summary measures of exposure<br>*Cross-sectional study* - Report numbers of outcome events or summary measures | Results, Tables 1-4, Fig 2, Fig. 3 | | |
| Main results | 16 | (a) Give unadjusted estimates and, if applicable, confounder-adjusted estimates and their precision (e.g., 95% confidence interval). Make clear which confounders were adjusted for and why they were included<br>(b) Report category boundaries when continuous variables were categorized<br>(c) If relevant, consider translating estimates of relative risk into absolute risk for a meaningful time period | Results, Tables 1-4, Fig. 2, Fig. 3 | | |
| Other analyses | 17 | Report other analyses done—e.g., analyses of subgroups and interactions, and sensitivity analyses | Results (3.3), Supplementary Tables 8-13 | | |
| **Discussion** | | | | | |
| Key results | 18 | Summarise key results with reference to study objectives | Abstract, Discussion (first paragraph, final paragraph) | | |
| Limitations | 19 | Discuss limitations of the study, taking into account sources of potential bias or imprecision. Discuss both direction and magnitude of any potential bias | Discussion | RECORD 19.1: Discuss the implications of using data that were not created or collected to answer the specific research question(s). Include discussion of misclassification bias, unmeasured confounding, missing data, and changing eligibility over time, as they pertain to the study being reported. | Discussion |
| Interpretation | 20 | Give a cautious overall interpretation of results considering objectives, limitations, multiplicity of analyses, results from similar studies, and other relevant evidence | Discussion | | |
| Generalisability | 21 | Discuss the generalisability (external validity) of the study results | Discussion | | |
| **Other Information** | | | | | |
| Funding | 22 | Give the source of funding and the role of the funders for the present study and, if applicable, for the original study on which the present article is based | Funding statement | | |

**Supplementary Table 2. Mild versus low inflammation in the period surrounding an adverse life event.** Demographic characteristics and other covariates of interest, including types of adverse events, comorbidities and hospital admissions within the last month before the record of adversity, in patients with mild leukocytosis (Cohort 1) and normal leukocyte counts (Cohort 2) in the period surrounding adverse life events, before and after matching. SMD: standardised mean difference.

| Cohort characteristics | | Cohort 1 (N = 37,442) and Cohort 2 (N = 109,504) before propensity score matching | | | | Cohort 1 (N = 36,772) and Cohort 2 (N = 36,772) after propensity score matching | | | |
|---|---|---|---|---|---|---|---|---|---|
| **Demographics** | | | | | | | | | |
| Cohort | | Mean ± SD | Patients | % of Cohort | SMD | Mean ± SD | Patients | % of Cohort | SMD |
| 1<br>2 | Age at Index | 44.9 +/- 19.5<br>46.8 +/- 19.5 | 37,442<br>109,504 | 100%<br>100% | 0.095 | 45.0 +/- 19.5<br>45.1 +/- 19.5 | 36,772<br>36,772 | 100%<br>100% | 0.003 |
| 1<br>2 | Female | | 16,413<br>51,811 | 43.8%<br>47.3% | 0.070 | | 16,219<br>16,343 | 44.1%<br>44.4% | 0.007 |
| 1<br>2 | Male | | 20,785<br>56,898 | 55.5%<br>52.0% | 0.071 | | 20,314<br>20,182 | 55.2%<br>54.9% | 0.007 |
| 1<br>2 | Black or African American | | 5,341<br>21,717 | 14.3%<br>19.8% | 0.148 | | 5,308<br>5,061 | 14.4%<br>13.8% | 0.019 |
| 1<br>2 | White | | 25,670<br>67,097 | 68.6%<br>61.3% | 0.153 | | 25,114<br>25,456 | 68.3%<br>69.2% | 0.020 |
| 1<br>2 | Asian | | 451<br>1,523 | 1.2%<br>1.4% | 0.016 | | 442<br>386 | 1.2%<br>1.0% | 0.014 |
| 1<br>2 | Native Hawaiian or Other Pacific Islander | | 90<br>255 | 0.2%<br>0.2% | 0.002 | | 87<br>66 | 0.2%<br>0.2% | 0.013 |
| 1<br>2 | American Indian or Alaska Native | | 442<br>1,321 | 1.2%<br>1.2% | 0.002 | | 436<br>415 | 1.2%<br>1.1% | 0.005 |
| 1<br>2 | Other Race | | 1,177<br>3,757 | 3.1%<br>3.4% | 0.016 | | 1,154<br>1,056 | 3.1%<br>2.9% | 0.016 |
| 1<br>2 | Unknown Race | | 4,271<br>13,834 | 11.4%<br>12.6% | 0.038 | | 4,231<br>4,332 | 11.5%<br>11.8% | 0.009 |
| 1<br>2 | Hispanic or Latino | | 3,935<br>11,858 | 10.5%<br>10.8% | 0.010 | | 3,870<br>3,724 | 10.5%<br>10.1% | 0.013 |
| 1<br>2 | Not Hispanic or Latino | | 26,985<br>75,796 | 72.1%<br>69.2% | 0.063 | | 26,434<br>26,562 | 71.9%<br>72.2% | 0.008 |
| 1<br>2 | Unknown Ethnicity | | 6,522<br>21,850 | 17.4%<br>20.0% | 0.065 | | 6,468<br>6,486 | 17.6%<br>17.6% | 0.001 |
| **Diagnosis** | | | | | | | | | |
| 1<br>2 | Mental and behavioral disorders due to psychoactive substance use | | 9,143<br>24,219 | 24.4%<br>22.1% | 0.055 | | 8,826<br>9,183 | 24.0%<br>25.0% | 0.023 |
| 1<br>2 | Mood [affective] disorders | | 3,413<br>9,624 | 9.1%<br>8.8% | 0.011 | | 3,286<br>3,842 | 8.9%<br>10.4% | 0.051 |

| | | | | | | | |
|---|---|---|---|---|---|---|---|
| 1<br>2 | Anxiety, dissociative, stress-related, somatoform and other nonpsychotic mental disorders | 3,207<br>8,492 | 8.6%<br>7.8% | 0.030 | 3,074<br>3,497 | 8.4%<br>9.5% | 0.040 |
| 1<br>2 | Schizophrenia, schizotypal, delusional, and other non-mood psychotic disorders | 772<br>2,335 | 2.1%<br>2.1% | 0.005 | 755<br>835 | 2.1%<br>2.3% | 0.015 |
| 1<br>2 | Disorders of adult personality and behavior | 283<br>750 | 0.8%<br>0.7% | 0.008 | 272<br>330 | 0.7%<br>0.9% | 0.018 |
| 1<br>2 | Intellectual Disabilities | 124<br>466 | 0.3%<br>0.4% | 0.015 | 123<br>172 | 0.3%<br>0.5% | 0.021 |
| 1<br>2 | Pervasive and specific developmental disorders | 102<br>319 | 0.3%<br>0.3% | 0.004 | 97<br>142 | 0.3%<br>0.4% | 0.022 |
| 1<br>2 | Polyneuropathies and other disorders of the peripheral nervous system | 398<br>1,162 | 1.1%<br>1.1% | <0.001 | 382<br>455 | 1.0%<br>1.2% | 0.019 |
| 1<br>2 | Nerve, nerve root and plexus disorders | 361<br>915 | 1.0%<br>0.8% | 0.014 | 344<br>338 | 0.9%<br>0.9% | 0.002 |
| 1<br>2 | Extrapyramidal and movement disorders | 510<br>1,810 | 1.4%<br>1.7% | 0.024 | 503<br>684 | 1.4%<br>1.9% | 0.039 |
| 1<br>2 | Inflammatory diseases of the central nervous system | 97<br>142 | 0.3%<br>0.1% | 0.029 | 90<br>64 | 0.2%<br>0.2% | 0.015 |
| 1<br>2 | Diseases of myoneural junction and muscle | 58<br>151 | 0.2%<br>0.1% | 0.004 | 53<br>60 | 0.1%<br>0.2% | 0.005 |
| 1<br>2 | Demyelinating diseases of the central nervous system | 133<br>357 | 0.4%<br>0.3% | 0.005 | 132<br>119 | 0.4%<br>0.3% | 0.006 |
| 1<br>2 | Neoplasms | 1,652<br>4,785 | 4.4%<br>4.4% | 0.002 | 1,614<br>1,599 | 4.4%<br>4.3% | 0.002 |
| 1<br>2 | Type 1 diabetes mellitus | 303<br>807 | 0.8%<br>0.7% | 0.008 | 297<br>319 | 0.8%<br>0.9% | 0.007 |
| 1<br>2 | Type 2 diabetes mellitus | 3,825<br>10,344 | 10.2%<br>9.4% | 0.026 | 3,748<br>3,691 | 10.2%<br>10.0% | 0.005 |
| 1<br>2 | Overweight and obesity | 2,146<br>4,262 | 5.7%<br>3.9% | 0.086 | 2,078<br>1,670 | 5.7%<br>4.5% | 0.050 |
| 1<br>2 | Hypertensive diseases | 7,674<br>20,093 | 20.5%<br>18.3% | 0.054 | 7,483<br>7,256 | 20.3%<br>19.7% | 0.015 |
| 1<br>2 | Other forms of heart disease | 7,149<br>17,518 | 19.1%<br>16.0% | 0.081 | 6,944<br>6,622 | 18.9%<br>18.0% | 0.023 |
| 1<br>2 | Ischemic heart diseases | 2,842<br>6,824 | 7.6%<br>6.2% | 0.054 | 2,761<br>2,627 | 7.5%<br>7.1% | 0.014 |
| 1<br>2 | Diseases of the blood and blood-forming organs and | 6,729<br>10,077 | 18.0%<br>9.2% | 0.258 | 6,148<br>5,906 | 16.7%<br>16.1% | 0.018 |

| | | | | | | | | |
|---|---|---|---|---|---|---|---|---|
| | certain disorders involving the immune mechanism | | | | | | | |
| 1<br>2 | Certain infectious and parasitic diseases | 2,802<br>6,454 | 7.5%<br>5.9% | 0.064 | | 2,676<br>2,642 | 7.3%<br>7.2% | 0.004 |
| 1<br>2 | Systemic connective tissue disorders | 210<br>676 | 0.6%<br>0.6% | 0.007 | | 207<br>176 | 0.6%<br>0.5% | 0.012 |
| 1<br>2 | Inflammatory polyarthropathies | 743<br>1,972 | 2.0%<br>1.8% | 0.013 | | 733<br>658 | 2.0%<br>1.8% | 0.015 |
| 1<br>2 | Diseases of the musculoskeletal system and connective tissue | 21,266<br>60,175 | 56.8%<br>55.0% | 0.037 | | 20,844<br>20,769 | 56.7%<br>56.5% | 0.004 |
| 1<br>2 | Certain disorders involving the immune mechanism | 145<br>508 | 0.4%<br>0.5% | 0.012 | | 145<br>112 | 0.4%<br>0.3% | 0.015 |
| 1<br>2 | Malignant neoplasms of lymphoid, hematopoietic and related tissue | 113<br>457 | 0.3%<br>0.4% | 0.019 | | 113<br>86 | 0.3%<br>0.2% | 0.014 |
| 1<br>2 | Persons with potential health hazards related to socioeconomic and psychosocial circumstances | 882<br>2,672 | 2.4%<br>2.4% | 0.006 | | 840<br>1,023 | 2.3%<br>2.8% | 0.032 |
| 1<br>2 | Nicotine dependence | 4,651<br>9,987 | 12.4%<br>9.1% | 0.107 | | 4,482<br>3,955 | 12.2%<br>10.8% | 0.045 |
| 1<br>2 | Encounter for examination and observation following transport accident | 13,166<br>37,350 | 35.2%<br>34.1% | 0.022 | | 12,931<br>12,536 | 35.2%<br>34.1% | 0.023 |
| 1<br>2 | Encounter for examination and observation following work accident | 697<br>3,335 | 1.9%<br>3.0% | 0.077 | | 696<br>682 | 1.9%<br>1.9% | 0.003 |
| 1<br>2 | Encounter for examination and observation following other accident | 25,154<br>69,971 | 67.2%<br>63.9% | 0.069 | | 24,646<br>24,947 | 67.0%<br>67.8% | 0.017 |
| 1<br>2 | Encounter for examination and observation following alleged rape | 590<br>3,545 | 1.6%<br>3.2% | 0.109 | | 590<br>578 | 1.6%<br>1.6% | 0.003 |
| 1<br>2 | Encounter for examination and observation following alleged physical abuse | 345<br>1,231 | 0.9%<br>1.1% | 0.020 | | 345<br>292 | 0.9%<br>0.8% | 0.016 |
| 1<br>2 | Problems related to social environment | 76<br>239 | 0.2%<br>0.2% | 0.003 | | 73<br>83 | 0.2%<br>0.2% | 0.006 |
| 1<br>2 | Other problems related to primary support group, including family circumstances | 113<br>292 | 0.3%<br>0.3% | 0.007 | | 110<br>119 | 0.3%<br>0.3% | 0.004 |

| | | | | | | | |
|---|---|---|---|---|---|---|---|
| 1<br>2 | Problems related to housing and economic circumstances | 462<br>1,447 | 1.2%<br>1.3% | 0.008 | 436<br>572 | 1.2%<br>1.6% | 0.032 |
| 1<br>2 | Problems related to employment and unemployment | 55<br>159 | 0.1%<br>0.1% | <0.001 | 47<br>73 | 0.1%<br>0.2% | 0.018 |
| **Procedure** | | | | | | | |
| 1<br>2 | Hospital Inpatient and Observation Care Services | 9,584<br>14,260 | 25.6%<br>13.0% | 0.323 | 8,944<br>9,016 | 24.3%<br>24.5% | 0.005 |
| 1<br>2 | Critical Care Services | 5,683<br>7,797 | 15.2%<br>7.1% | 0.258 | 5,108<br>4,998 | 13.9%<br>13.6% | 0.009 |

**Supplementary Table 3. Mild versus low inflammation in the period surrounding a record of a psychosocial stressor.** Demographic characteristics and other covariates of interest, including types of adverse events, comorbidities and hospital admissions within the last month before the record of adversity, in patients with mild leukocytosis (Cohort 1) and normal leukocyte counts (Cohort 2) in the period surrounding a record of a psychosocial stressor, before and after matching. SMD: standardised mean difference.

| Cohort characteristics | | Cohort 1 (N = 88,620) and Cohort 2 (N = 365,381) before propensity score matching | | | | Cohort 1 (N = 87,936) and Cohort 2 (N = 87,936) after propensity score matching | | | |
|---|---|---|---|---|---|---|---|---|---|
| **Demographics** | | | | | | | | | |
| Cohort | | Mean ± SD | Patients | % of Cohort | SMD | Mean ± SD | Patients | % of Cohort | SMD |
| 1<br>2 | Age at Index | 45.8 +/- 17.1<br>45.9 +/- 17.5 | 88,620<br>365,381 | 100%<br>100% | 0.007 | 45.8 +/- 17.1<br>45.5 +/- 17.1 | 87,936<br>87,936 | 100%<br>100% | 0.018 |
| 1<br>2 | Female | | 42,374<br>182,234 | 47.8%<br>49.9% | 0.041 | | 42,109<br>42,390 | 47.9%<br>48.2% | 0.006 |
| 1<br>2 | Male | | 42,161<br>162,577 | 47.6%<br>44.5% | 0.062 | | 41,753<br>41,781 | 47.5%<br>47.5% | 0.001 |
| 1<br>2 | Black or African American | | 15,766<br>81,332 | 17.8%<br>22.3% | 0.112 | | 15,716<br>15,331 | 17.9%<br>17.4% | 0.011 |
| 1<br>2 | White | | 53,727<br>203,937 | 60.6%<br>55.8% | 0.098 | | 53,227<br>54,233 | 60.5%<br>61.7% | 0.023 |
| 1<br>2 | Asian | | 2,174<br>9,463 | 2.5%<br>2.6% | 0.009 | | 2,153<br>1,894 | 2.4%<br>2.2% | 0.020 |
| 1<br>2 | Native Hawaiian or Other Pacific Islander | | 1,189<br>3,148 | 1.3%<br>0.9% | 0.046 | | 1,151<br>1,114 | 1.3%<br>1.3% | 0.004 |
| 1<br>2 | American Indian or Alaska Native | | 1,068<br>3,515 | 1.2%<br>1.0% | 0.023 | | 1,056<br>996 | 1.2%<br>1.1% | 0.006 |
| 1<br>2 | Other Race | | 3,027<br>12,397 | 3.4%<br>3.4% | 0.001 | | 3,006<br>2,952 | 3.4%<br>3.4% | 0.003 |
| 1<br>2 | Unknown Race | | 11,669<br>51,589 | 13.2%<br>14.1% | 0.028 | | 11,627<br>11,416 | 13.2%<br>13.0% | 0.007 |
| 1<br>2 | Hispanic or Latino | | 9,144<br>35,090 | 10.3%<br>9.6% | 0.024 | | 9,061<br>8,906 | 10.3%<br>10.1% | 0.006 |
| 1<br>2 | Not Hispanic or Latino | | 54,267<br>222,289 | 61.2%<br>60.8% | 0.008 | | 53,793<br>53,676 | 61.2%<br>61.0% | 0.003 |
| 1<br>2 | Unknown Ethnicity | | 25,209<br>108,002 | 28.4%<br>29.6% | 0.025 | | 25,082<br>25,354 | 28.5%<br>28.8% | 0.007 |
| **Diagnosis** | | | | | | | | | |
| 1<br>2 | Mental and behavioral disorders due to psychoactive substance use | | 45,862<br>152,726 | 51.8%<br>41.8% | 0.200 | | 45,444<br>42,289 | 51.7%<br>48.1% | 0.072 |

| | | | | | | | |
|---|---|---|---|---|---|---|---|
| 1 | Mood [affective] disorders | 33,995 | 38.4% | 0.015 | 33,732 | 38.4% | 0.042 |
| 2 | | 142,849 | 39.1% | | 35,522 | 40.4% | |
| 1 | Anxiety, dissociative, stress-related, somatoform and other nonpsychotic mental disorders | 30,568 | 34.5% | 0.019 | 30,312 | 34.5% | 0.032 |
| 2 | | 129,405 | 35.4% | | 31,645 | 36.0% | |
| 1 | Schizophrenia, schizotypal, delusional, and other non-mood psychotic disorders | 10,044 | 11.3% | 0.001 | 9,999 | 11.4% | 0.012 |
| 2 | | 41,505 | 11.4% | | 10,341 | 11.8% | |
| 1 | Disorders of adult personality and behavior | 4,516 | 5.1% | 0.004 | 4,477 | 5.1% | 0.011 |
| 2 | | 18,284 | 5.0% | | 4,696 | 5.3% | |
| 1 | Intellectual Disabilities | 766 | 0.9% | 0.001 | 756 | 0.9% | 0.007 |
| 2 | | 3,182 | 0.9% | | 817 | 0.9% | |
| 1 | Pervasive and specific developmental disorders | 805 | 0.9% | 0.021 | 793 | 0.9% | 0.019 |
| 2 | | 4,083 | 1.1% | | 959 | 1.1% | |
| 1 | Polyneuropathies and other disorders of the peripheral nervous system | 2,575 | 2.9% | 0.043 | 2,532 | 2.9% | 0.005 |
| 2 | | 8,156 | 2.2% | | 2,457 | 2.8% | |
| 1 | Nerve, nerve root and plexus disorders | 1,451 | 1.6% | 0.028 | 1,422 | 1.6% | 0.013 |
| 2 | | 4,750 | 1.3% | | 1,277 | 1.5% | |
| 1 | Extrapyramidal and movement disorders | 2,319 | 2.6% | 0.029 | 2,286 | 2.6% | <0.001 |
| 2 | | 7,920 | 2.2% | | 2,289 | 2.6% | |
| 1 | Inflammatory diseases of the central nervous system | 539 | 0.6% | 0.056 | 529 | 0.6% | 0.022 |
| 2 | | 884 | 0.2% | | 390 | 0.4% | |
| 1 | Diseases of myoneural junction and muscle | 367 | 0.4% | 0.027 | 360 | 0.4% | 0.012 |
| 2 | | 946 | 0.3% | | 298 | 0.3% | |
| 1 | Demyelinating diseases of the central nervous system | 508 | 0.6% | 0.011 | 501 | 0.6% | 0.008 |
| 2 | | 1,809 | 0.5% | | 449 | 0.5% | |
| 1 | Neoplasms | 8,346 | 9.4% | 0.075 | 8,246 | 9.4% | 0.048 |
| 2 | | 26,860 | 7.4% | | 7,052 | 8.0% | |
| 1 | Type 1 diabetes mellitus | 1,884 | 2.1% | 0.063 | 1,865 | 2.1% | 0.036 |
| 2 | | 4,797 | 1.3% | | 1,432 | 1.6% | |
| 1 | Type 2 diabetes mellitus | 18,758 | 21.2% | 0.150 | 18,542 | 21.1% | 0.093 |
| 2 | | 56,168 | 15.4% | | 15,107 | 17.2% | |
| 1 | Overweight and obesity | 15,755 | 17.8% | 0.166 | 15,573 | 17.7% | 0.093 |
| 2 | | 43,478 | 11.9% | | 12,633 | 14.4% | |
| 1 | Hypertensive diseases | 35,519 | 40.1% | 0.145 | 35,123 | 39.9% | 0.092 |
| 2 | | 120,915 | 33.1% | | 31,203 | 35.5% | |
| 1 | Other forms of heart disease | 20,509 | 23.1% | 0.212 | 20,129 | 22.9% | 0.083 |
| 2 | | 54,283 | 14.9% | | 17,139 | 19.5% | |
| 1 | Ischemic heart diseases | 11,751 | 13.3% | 0.158 | 11,560 | 13.1% | 0.075 |
| 2 | | 30,550 | 8.4% | | 9,432 | 10.7% | |

| | | | | | | | |
|---|---|---|---|---|---|---|---|
| 1<br>2 | Diseases of the blood and blood-forming organs and certain disorders involving the immune mechanism | 28,308<br>60,791 | 31.9%<br>16.6% | 0.363 | 27,651<br>27,419 | 31.4%<br>31.2% | 0.006 |
| 1<br>2 | Certain infectious and parasitic diseases | 22,083<br>55,156 | 24.9%<br>15.1% | 0.247 | 21,627<br>21,541 | 24.6%<br>24.5% | 0.002 |
| 1<br>2 | Systemic connective tissue disorders | 1,086<br>4,048 | 1.2%<br>1.1% | 0.011 | 1,068<br>886 | 1.2%<br>1.0% | 0.020 |
| 1<br>2 | Inflammatory polyarthropathies | 3,585<br>11,356 | 4.0%<br>3.1% | 0.050 | 3,515<br>3,203 | 4.0%<br>3.6% | 0.019 |
| 1<br>2 | Diseases of the musculoskeletal system and connective tissue | 32,573<br>112,104 | 36.8%<br>30.7% | 0.129 | 32,187<br>28,674 | 36.6%<br>32.6% | 0.084 |
| 1<br>2 | Certain disorders involving the immune mechanism | 1,264<br>3,931 | 1.4%<br>1.1% | 0.032 | 1,253<br>1,057 | 1.4%<br>1.2% | 0.020 |
| 1<br>2 | Malignant neoplasms of lymphoid, hematopoietic and related tissue | 823<br>3,065 | 0.9%<br>0.8% | 0.010 | 819<br>721 | 0.9%<br>0.8% | 0.012 |
| 1<br>2 | Nicotine dependence | 33,431<br>100,330 | 37.7%<br>27.5% | 0.220 | 33,137<br>28,946 | 37.7%<br>32.9% | 0.100 |
| 1<br>2 | Encounter for examination and observation following transport accident | 147<br>263 | 0.2%<br>0.1% | 0.027 | 144<br>67 | 0.2%<br>0.1% | 0.025 |
| 1<br>2 | Encounter for examination and observation following work accident | 10<br>10 | 0.0%<br>0.0% | 0.010 | 10<br>10 | 0.0%<br>0.0% | <0.001 |
| 1<br>2 | Encounter for examination and observation following other accident | 526<br>1,241 | 0.6%<br>0.3% | 0.037 | 512<br>428 | 0.6%<br>0.5% | 0.013 |
| 1<br>2 | Encounter for examination and observation following alleged rape | 29<br>134 | 0.0%<br>0.0% | 0.002 | 28<br>36 | 0.0%<br>0.0% | 0.005 |
| 1<br>2 | Encounter for examination and observation following alleged physical abuse | 18<br>85 | 0.0%<br>0.0% | 0.002 | 18<br>23 | 0.0%<br>0.0% | 0.004 |
| 1<br>2 | Problems related to social environment | 14,686<br>55,908 | 16.6%<br>15.3% | 0.035 | 14,548<br>14,320 | 16.5%<br>16.3% | 0.029 |
| 1<br>2 | Other problems related to primary support group, including family circumstances | 21,123<br>113,040 | 23.8%<br>30.9% | 0.160 | 21,053<br>19,957 | 23.9%<br>22.7% | 0.021 |
| 1<br>2 | Problems related to housing and economic circumstances | 48,065<br>174,795 | 54.2%<br>47.8% | 0.128 | 47,616<br>48,539 | 54.1%<br>55.2% | 0.016 |

| | | | | | | | |
|---|---|---|---|---|---|---|---|
| 1<br>2 | Problems related to employment and unemployment | 13,113<br>59,063 | 14.8%<br>16.2% | 0.038 | 13,037<br>12,547 | 14.8%<br>14.3% | 0.066 |
| 1<br>2 | Major depressive disorder, recurrent | 7,270<br>38,362 | 8.2%<br>10.5% | 0.079 | 7,226<br>8,899 | 8.2%<br>10.1% | 0.029 |
| **Procedure** | | | | | | | |
| 1<br>2 | Hospital Inpatient and Observation Care Services | 29,929<br>68,655 | 33.8%<br>18.8% | 0.345 | 29,281<br>28,969 | 33.3%<br>32.9% | 0.008 |
| 1<br>2 | Critical Care Services | 9,789<br>12,488 | 11.0%<br>3.4% | 0.298 | 9,118<br>8,434 | 10.4%<br>9.6% | 0.026 |

**Supplementary Table 4. Dynamic versus low inflammation in the period surrounding an adverse life event.** Demographic characteristics and other covariates of interest, including types of adverse events, comorbidities and hospital admissions within the last month before the record of adversity, in patients with dynamic changes in (Cohort 1) and persistent normal leukocyte counts (Cohort 2) in the period surrounding a record of a psychosocial stressor, before and after matching. SMD: standardised mean difference.

| Cohort characteristics | | Cohort 1 (N = 13,425) and Cohort 2 (N = 33,140) before propensity score matching | | | | Cohort 1 (N = 12,303) and Cohort 2 (N = 12,303) after propensity score matching | | | |
|---|---|---|---|---|---|---|---|---|---|
| **Demographics** | | | | | | | | | |
| Cohort | | Mean ± SD | Patients | % of Cohort | SMD | Mean ± SD | Patients | % of Cohort | SMD |
| 1<br>2 | Age at Index | 51.9 +/- 19.2<br>53.1 +/- 18.5 | 13,425<br>33,140 | 100%<br>100% | 0.066 | 52.3 +/- 19.2<br>52.9 +/- 18.7 | 12,303<br>12,303 | 100%<br>100% | 0.031 |
| 1<br>2 | Female | | 6,176<br>17,493 | 46.0%<br>52.8% | 0.136 | | 5,832<br>5,883 | 47.4%<br>47.8% | 0.008 |
| 1<br>2 | Male | | 7,165<br>15,331 | 53.4%<br>46.3% | 0.143 | | 6,388<br>6,342 | 51.9%<br>51.5% | 0.007 |
| 1<br>2 | Black or African American | | 1,721<br>6,144 | 12.8%<br>18.5% | 0.158 | | 1,649<br>1,561 | 13.4%<br>12.7% | 0.021 |
| 1<br>2 | White | | 10,047<br>21,244 | 74.8%<br>64.1% | 0.235 | | 9,074<br>9,236 | 73.8%<br>75.1% | 0.030 |
| 1<br>2 | Asian | | 138<br>376 | 1.0%<br>1.1% | 0.010 | | 131<br>116 | 1.1%<br>0.9% | 0.012 |
| 1<br>2 | Native Hawaiian or Other Pacific Islander | | 24<br>69 | 0.2%<br>0.2% | 0.007 | | 21<br>18 | 0.2%<br>0.1% | 0.006 |
| 1<br>2 | American Indian or Alaska Native | | 101<br>348 | 0.8%<br>1.1% | 0.032 | | 97<br>81 | 0.8%<br>0.7% | 0.015 |
| 1<br>2 | Other Race | | 340<br>893 | 2.5%<br>2.7% | 0.010 | | 306<br>275 | 2.5%<br>2.2% | 0.017 |
| 1<br>2 | Unknown Race | | 1,054<br>4,066 | 7.9%<br>12.3% | 0.147 | | 1,025<br>1,016 | 8.3%<br>8.3% | 0.003 |
| 1<br>2 | Hispanic or Latino | | 1,014<br>2,949 | 7.6%<br>8.9% | 0.049 | | 940<br>833 | 7.6%<br>6.8% | 0.034 |
| 1<br>2 | Not Hispanic or Latino | | 10,442<br>23,279 | 77.8%<br>70.2% | 0.172 | | 9,470<br>9,617 | 77.0%<br>78.2% | 0.029 |
| 1<br>2 | Unknown Ethnicity | | 1,969<br>6,912 | 14.7%<br>20.9% | 0.163 | | 1,893<br>1,853 | 15.4%<br>15.1% | 0.009 |
| **Diagnosis** | | | | | | | | | |
| 1<br>2 | Mental and behavioral disorders due to psychoactive substance use | | 3,635<br>7,656 | 27.1%<br>23.1% | 0.092 | | 3,190<br>3,368 | 25.9%<br>27.4% | 0.033 |

| | | | | | | | |
|---|---|---|---|---|---|---|---|
| 1<br>2 | Mood [affective] disorders | 2,051<br>5,103 | 15.3%<br>15.4% | 0.003 | 1,828<br>2,189 | 14.9%<br>17.8% | 0.079 |
| 1<br>2 | Anxiety, dissociative, stress-related, somatoform and other nonpsychotic mental disorders | 1,895<br>4,248 | 14.1%<br>12.8% | 0.038 | 1,689<br>1,891 | 13.7%<br>15.4% | 0.047 |
| 1<br>2 | Schizophrenia, schizotypal, delusional, and other non-mood psychotic disorders | 481<br>1,174 | 3.6%<br>3.5% | 0.002 | 424<br>467 | 3.4%<br>3.8% | 0.019 |
| 1<br>2 | Disorders of adult personality and behavior | 185<br>421 | 1.4%<br>1.3% | 0.009 | 167<br>174 | 1.4%<br>1.4% | 0.005 |
| 1<br>2 | Intellectual Disabilities | 77<br>267 | 0.6%<br>0.8% | 0.028 | 73<br>113 | 0.6%<br>0.9% | 0.038 |
| 1<br>2 | Pervasive and specific developmental disorders | 54<br>149 | 0.4%<br>0.4% | 0.007 | 48<br>74 | 0.4%<br>0.6% | 0.030 |
| 1<br>2 | Polyneuropathies and other disorders of the peripheral nervous system | 319<br>724 | 2.4%<br>2.2% | 0.013 | 292<br>336 | 2.4%<br>2.7% | 0.023 |
| 1<br>2 | Nerve, nerve root and plexus disorders | 223<br>476 | 1.7%<br>1.4% | 0.018 | 194<br>196 | 1.6%<br>1.6% | 0.001 |
| 1<br>2 | Extrapyramidal and movement disorders | 360<br>1,015 | 2.7%<br>3.1% | 0.023 | 317<br>444 | 2.6%<br>3.6% | 0.060 |
| 1<br>2 | Inflammatory diseases of the central nervous system | 63<br>89 | 0.5%<br>0.3% | 0.033 | 51<br>46 | 0.4%<br>0.4% | 0.006 |
| 1<br>2 | Diseases of myoneural junction and muscle | 43<br>84 | 0.3%<br>0.3% | 0.012 | 38<br>36 | 0.3%<br>0.3% | 0.003 |
| 1<br>2 | Demyelinating diseases of the central nervous system | 73<br>177 | 0.5%<br>0.5% | 0.001 | 70<br>61 | 0.6%<br>0.5% | 0.010 |
| 1<br>2 | Neoplasms | 1,145<br>2,957 | 8.5%<br>8.9% | 0.014 | 1,067<br>1,112 | 8.7%<br>9.0% | 0.013 |
| 1<br>2 | Type 1 diabetes mellitus | 178<br>411 | 1.3%<br>1.2% | 0.008 | 165<br>188 | 1.3%<br>1.5% | 0.016 |
| 1<br>2 | Type 2 diabetes mellitus | 2,290<br>5,557 | 17.1%<br>16.8% | 0.008 | 2,098<br>2,269 | 17.1%<br>18.4% | 0.036 |
| 1<br>2 | Overweight and obesity | 1,199<br>2,328 | 8.9%<br>7.0% | 0.070 | 1,071<br>1,037 | 8.7%<br>8.4% | 0.010 |
| 1<br>2 | Hypertensive diseases | 4,428<br>10,039 | 33.0%<br>30.3% | 0.058 | 4,009<br>4,202 | 32.6%<br>34.2% | 0.033 |
| 1<br>2 | Other forms of heart disease | 4,323<br>8,602 | 32.2%<br>26.0% | 0.138 | 3,877<br>3,799 | 31.5%<br>30.9% | 0.014 |
| 1<br>2 | Ischemic heart diseases | 1,816<br>3,651 | 13.5%<br>11.0% | 0.077 | 1,628<br>1,638 | 13.2%<br>13.3% | 0.002 |

| | | | | | | | |
|---|---|---|---|---|---|---|---|
| 1<br>2 | Cerebrovascular diseases | 1,916<br>3,309 | 14.3%<br>10.0% | 0.132 | 1,660<br>1,558 | 13.5%<br>12.7% | 0.025 |
| 1<br>2 | Diseases of the blood and blood-forming organs and certain disorders involving the immune mechanism | 4,281<br>5,808 | 31.9%<br>17.5% | 0.338 | 3,507<br>3,469 | 28.5%<br>28.2% | 0.007 |
| 1<br>2 | Certain infectious and parasitic diseases | 2,028<br>3,811 | 15.1%<br>11.5% | 0.106 | 1,781<br>1,852 | 14.5%<br>15.1% | 0.016 |
| 1<br>2 | Systemic connective tissue disorders | 142<br>403 | 1.1%<br>1.2% | 0.015 | 134<br>126 | 1.1%<br>1.0% | 0.006 |
| 1<br>2 | Inflammatory polyarthropathies | 463<br>1,098 | 3.4%<br>3.3% | 0.008 | 418<br>388 | 3.4%<br>3.2% | 0.014 |
| 1<br>2 | Diseases of the musculoskeletal system and connective tissue | 8,593<br>19,894 | 64.0%<br>60.0% | 0.082 | 7,830<br>7,929 | 63.6%<br>64.4% | 0.017 |
| 1<br>2 | Certain disorders involving the immune mechanism | 111<br>394 | 0.8%<br>1.2% | 0.036 | 109<br>100 | 0.9%<br>0.8% | 0.008 |
| 1<br>2 | Malignant neoplasms of lymphoid, hematopoietic and related tissue | 101<br>393 | 0.8%<br>1.2% | 0.044 | 97<br>85 | 0.8%<br>0.7% | 0.011 |
| 1<br>2 | Nicotine dependence | 2,002<br>3,839 | 14.9%<br>11.6% | 0.098 | 1,755<br>1,735 | 14.3%<br>14.1% | 0.005 |
| 1<br>2 | Encounter for examination and observation following transport accident | 3,511<br>6,627 | 26.2%<br>20.0% | 0.146 | 3,071<br>2,968 | 25.0%<br>24.1% | 0.019 |
| 1<br>2 | Encounter for examination and observation following work accident | 179<br>1,630 | 1.3%<br>4.9% | 0.207 | 179<br>178 | 1.5%<br>1.4% | 0.001 |
| 1<br>2 | Encounter for examination and observation following other accident | 10,280<br>24,295 | 76.6%<br>73.3% | 0.075 | 9,450<br>9,572 | 76.8%<br>77.8% | 0.024 |
| 1<br>2 | Encounter for examination and observation following alleged rape | 165<br>1,165 | 1.2%<br>3.5% | 0.151 | 164<br>157 | 1.3%<br>1.3% | 0.005 |
| 1<br>2 | Encounter for examination and observation following alleged physical abuse | 115<br>415 | 0.9%<br>1.3% | 0.039 | 113<br>93 | 0.9%<br>0.8% | 0.018 |
| 1<br>2 | Problems related to social environment | 57<br>127 | 0.4%<br>0.4% | 0.007 | 54<br>50 | 0.4%<br>0.4% | 0.005 |
| 1<br>2 | Other problems related to primary support group, including family circumstances | 69<br>156 | 0.5%<br>0.5% | 0.006 | 63<br>52 | 0.5%<br>0.4% | 0.013 |

| | | | | | | | |
|---|---|---|---|---|---|---|---|
| 1<br>2 | Problems related to housing and economic circumstances | 283<br>871 | 2.1%<br>2.6% | 0.034 | 233<br>384 | 1.9%<br>3.1% | 0.079 |
| 1<br>2 | Problems related to employment and unemployment | 32<br>77 | 0.2%<br>0.2% | 0.001 | 27<br>38 | 0.2%<br>0.3% | 0.017 |
| 1<br>2 | Major depressive disorder, recurrent | 352<br>868 | 2.6%<br>2.6% | <0.001 | 312<br>387 | 2.5%<br>3.1% | 0.037 |
| **Procedure** | | | | | | | |
| 1<br>2 | Hospital Inpatient and Observation Care Services | 5,265<br>6,400 | 39.2%<br>19.3% | 0.448 | 4,334<br>4,399 | 35.2%<br>35.8% | 0.011 |
| 1<br>2 | Critical Care Services | 3,052<br>2,257 | 22.7%<br>6.8% | 0.461 | 2,060<br>1,994 | 16.7%<br>16.2% | 0.014 |

**Supplementary Table 5. Dynamic versus high inflammation in the period surrounding an adverse life event.** Demographic characteristics and other covariates of interest, including types of adverse events, comorbidities and hospital admissions within the last month before the record of adversity, in patients with dynamic changes in (Cohort 1) and persistent high leukocyte counts (Cohort 2) in the period surrounding a record of a psychosocial stressor, before and after matching. SMD: standardised mean difference.

| Cohort characteristics | | Cohort 1 (N = 13,425) and Cohort 2 (N = 16,393) before propensity score matching | | | | Cohort 1 (N = 12,874) and Cohort 2 (N = 12,874) after propensity score matching | | | |
|---|---|---|---|---|---|---|---|---|---|
| **Demographics** | | | | | | | | | |
| Cohort | | Mean ± SD | Patients | % of Cohort | SMD | Mean ± SD | Patients | % of Cohort | SMD |
| 1<br>2 | Age at Index | 51.9 +/- 19.2<br>49.7 +/- 19.1 | 13,425<br>16,393 | 100%<br>100% | 0.114 | 51.3 +/- 19.2<br>51.3 +/- 19.1 | 12,874<br>12,874 | 100%<br>100% | 0.003 |
| 1<br>2 | Female | | 6,176<br>6,999 | 46.0%<br>42.7% | 0.067 | | 5,823<br>5,836 | 45.2%<br>45.3% | 0.002 |
| 1<br>2 | Male | | 7,165<br>9,295 | 53.4%<br>56.7% | 0.067 | | 6,971<br>6,954 | 54.1%<br>54.0% | 0.003 |
| 1<br>2 | Black or African American | | 1,721<br>2,455 | 12.8%<br>15.0% | 0.062 | | 1,699<br>1,687 | 13.2%<br>13.1% | 0.003 |
| 1<br>2 | White | | 10,047<br>11,543 | 74.8%<br>70.4% | 0.099 | | 9,560<br>9,579 | 74.3%<br>74.4% | 0.003 |
| 1<br>2 | Asian | | 138<br>147 | 1.0%<br>0.9% | 0.013 | | 125<br>124 | 1.0%<br>1.0% | 0.001 |
| 1<br>2 | Native Hawaiian or Other Pacific Islander | | 24<br>25 | 0.2%<br>0.2% | 0.006 | | 22<br>21 | 0.2%<br>0.2% | 0.002 |
| 1<br>2 | American Indian or Alaska Native | | 101<br>126 | 0.8%<br>0.8% | 0.002 | | 97<br>93 | 0.8%<br>0.7% | 0.004 |
| 1<br>2 | Other Race | | 340<br>392 | 2.5%<br>2.4% | 0.009 | | 322<br>306 | 2.5%<br>2.4% | 0.008 |
| 1<br>2 | Unknown Race | | 1,054<br>1,705 | 7.9%<br>10.4% | 0.089 | | 1,049<br>1,064 | 8.1%<br>8.3% | 0.004 |
| 1<br>2 | Hispanic or Latino | | 1,014<br>1,220 | 7.6%<br>7.4% | 0.004 | | 968<br>943 | 7.5%<br>7.3% | 0.007 |
| 1<br>2 | Not Hispanic or Latino | | 10,442<br>12,349 | 77.8%<br>75.3% | 0.058 | | 9,962<br>9,984 | 77.4%<br>77.6% | 0.004 |
| 1<br>2 | Unknown Ethnicity | | 1,969<br>2,824 | 14.7%<br>17.2% | 0.070 | | 1,944<br>1,947 | 15.1%<br>15.1% | 0.001 |
| **Diagnosis** | | | | | | | | | |
| 1<br>2 | Mental and behavioral disorders due to psychoactive substance use | | 3,635<br>4,785 | 27.1%<br>29.2% | 0.047 | | 3,535<br>3,543 | 27.5%<br>27.5% | 0.001 |

| | | | | | | | |
|---|---|---|---|---|---|---|---|
| 1<br>2 | Mood [affective] disorders | 2,051<br>2,521 | 15.3%<br>15.4% | 0.003 | 1,974<br>1,938 | 15.3%<br>15.1% | 0.008 |
| 1<br>2 | Anxiety, dissociative, stress-related, somatoform and other nonpsychotic mental disorders | 1,895<br>2,445 | 14.1%<br>14.9% | 0.023 | 1,820<br>1,790 | 14.1%<br>13.9% | 0.007 |
| 1<br>2 | Schizophrenia, schizotypal, delusional, and other non-mood psychotic disorders | 481<br>553 | 3.6%<br>3.4% | 0.011 | 469<br>409 | 3.6%<br>3.2% | 0.026 |
| 1<br>2 | Disorders of adult personality and behavior | 185<br>235 | 1.4%<br>1.4% | 0.005 | 183<br>181 | 1.4%<br>1.4% | 0.001 |
| 1<br>2 | Intellectual Disabilities | 77<br>90 | 0.6%<br>0.5% | 0.003 | 76<br>71 | 0.6%<br>0.6% | 0.005 |
| 1<br>2 | Pervasive and specific developmental disorders | 54<br>79 | 0.4%<br>0.5% | 0.012 | 53<br>55 | 0.4%<br>0.4% | 0.002 |
| 1<br>2 | Polyneuropathies and other disorders of the peripheral nervous system | 319<br>394 | 2.4%<br>2.4% | 0.002 | 304<br>285 | 2.4%<br>2.2% | 0.010 |
| 1<br>2 | Nerve, nerve root and plexus disorders | 223<br>330 | 1.7%<br>2.0% | 0.026 | 215<br>208 | 1.7%<br>1.6% | 0.004 |
| 1<br>2 | Extrapyramidal and movement disorders | 360<br>381 | 2.7%<br>2.3% | 0.023 | 342<br>296 | 2.7%<br>2.3% | 0.023 |
| 1<br>2 | Inflammatory diseases of the central nervous system | 63<br>194 | 0.5%<br>1.2% | 0.079 | 62<br>117 | 0.5%<br>0.9% | 0.051 |
| 1<br>2 | Diseases of myoneural junction and muscle | 43<br>93 | 0.3%<br>0.6% | 0.037 | 43<br>68 | 0.3%<br>0.5% | 0.030 |
| 1<br>2 | Demyelinating diseases of the central nervous system | 73<br>91 | 0.5%<br>0.6% | 0.002 | 69<br>74 | 0.5%<br>0.6% | 0.005 |
| 1<br>2 | Neoplasms | 1,145<br>1,561 | 8.5%<br>9.5% | 0.035 | 1,086<br>1,210 | 8.4%<br>9.4% | 0.034 |
| 1<br>2 | Type 1 diabetes mellitus | 178<br>251 | 1.3%<br>1.5% | 0.017 | 175<br>203 | 1.4%<br>1.6% | 0.018 |
| 1<br>2 | Type 2 diabetes mellitus | 2,290<br>2,883 | 17.1%<br>17.6% | 0.014 | 2,172<br>2,336 | 16.9%<br>18.1% | 0.034 |
| 1<br>2 | Overweight and obesity | 1,199<br>1,797 | 8.9%<br>11.0% | 0.068 | 1,164<br>1,345 | 9.0%<br>10.4% | 0.047 |
| 1<br>2 | Hypertensive diseases | 4,428<br>5,604 | 33.0%<br>34.2% | 0.025 | 4,214<br>4,371 | 32.7%<br>34.0% | 0.026 |
| 1<br>2 | Other forms of heart disease | 4,323<br>5,764 | 32.2%<br>35.2% | 0.063 | 4,118<br>4,451 | 32.0%<br>34.6% | 0.055 |
| 1<br>2 | Ischemic heart diseases | 1,816<br>2,303 | 13.5%<br>14.0% | 0.015 | 1,730<br>1,823 | 13.4%<br>14.2% | 0.021 |

| | | | | | | | |
|---|---|---|---|---|---|---|---|
| 1<br>2 | Diseases of the blood and blood-forming organs and certain disorders involving the immune mechanism | 4,281<br>6,782 | 31.9%<br>41.4% | 0.198 | 4,278<br>4,316 | 33.2%<br>33.5% | 0.006 |
| 1<br>2 | Certain infectious and parasitic diseases | 2,028<br>3,547 | 15.1%<br>21.6% | 0.169 | 2,027<br>1,990 | 15.7%<br>15.5% | 0.008 |
| 1<br>2 | Systemic connective tissue disorders | 142<br>171 | 1.1%<br>1.0% | 0.001 | 138<br>132 | 1.1%<br>1.0% | 0.005 |
| 1<br>2 | Inflammatory polyarthropathies | 463<br>587 | 3.4%<br>3.6% | 0.007 | 445<br>440 | 3.5%<br>3.4% | 0.002 |
| 1<br>2 | Diseases of the musculoskeletal system and connective tissue | 8,593<br>9,995 | 64.0%<br>61.0% | 0.063 | 8,206<br>7,912 | 63.7%<br>61.5% | 0.047 |
| 1<br>2 | Certain disorders involving the immune mechanism | 111<br>167 | 0.8%<br>1.0% | 0.020 | 111<br>109 | 0.9%<br>0.8% | 0.002 |
| 1<br>2 | Malignant neoplasms of lymphoid, hematopoietic and related tissue | 101<br>155 | 0.8%<br>0.9% | 0.021 | 101<br>105 | 0.8%<br>0.8% | 0.003 |
| 1<br>2 | Nicotine dependence | 2,002<br>2,823 | 14.9%<br>17.2% | 0.063 | 1,943<br>2,093 | 15.1%<br>16.3% | 0.032 |
| 1<br>2 | Encounter for examination and observation following transport accident | 3,511<br>4,391 | 26.2%<br>26.8% | 0.014 | 3,412<br>3,335 | 26.5%<br>25.9% | 0.014 |
| 1<br>2 | Encounter for examination and observation following work accident | 179<br>390 | 1.3%<br>2.4% | 0.078 | 179<br>187 | 1.4%<br>1.5% | 0.005 |
| 1<br>2 | Encounter for examination and observation following other accident | 10,280<br>12,326 | 76.6%<br>75.2% | 0.032 | 9,819<br>9,873 | 76.3%<br>76.7% | 0.010 |
| 1<br>2 | Encounter for examination and observation following alleged rape | 165<br>187 | 1.2%<br>1.1% | 0.008 | 161<br>164 | 1.3%<br>1.3% | 0.002 |
| 1<br>2 | Encounter for examination and observation following alleged physical abuse | 115<br>115 | 0.9%<br>0.7% | 0.018 | 101<br>97 | 0.8%<br>0.8% | 0.004 |
| 1<br>2 | Problems related to social environment | 57<br>59 | 0.4%<br>0.4% | 0.010 | 55<br>45 | 0.4%<br>0.3% | 0.012 |
| 1<br>2 | Other problems related to primary support group, including family circumstances | 69<br>69 | 0.5%<br>0.4% | 0.014 | 66<br>49 | 0.5%<br>0.4% | 0.020 |
| 1<br>2 | Problems related to housing and economic circumstances | 283<br>373 | 2.1%<br>2.3% | 0.011 | 279<br>252 | 2.2%<br>2.0% | 0.015 |

| | | | | | | | | |
|---|---|---|---|---|---|---|---|---|
| 1<br>2 | Problems related to employment and unemployment | 32<br>43 | 0.2%<br>0.3% | 0.005 | | 32<br>27 | 0.2%<br>0.2% | 0.008 |
| 1<br>2 | Major depressive disorder, recurrent | 352<br>356 | 2.6%<br>2.2% | 0.029 | | 337<br>286 | 2.6%<br>2.2% | 0.026 |
| **Procedure** | | | | | | | | |
| 1<br>2 | Hospital Inpatient and Observation Care Services | 5,265<br>6,458 | 39.2%<br>39.4% | 0.004 | | 4,997<br>4,995 | 38.8%<br>38.8% | <0.001 |
| 1<br>2 | Critical Care Services | 3,052<br>4,884 | 22.7%<br>29.8% | 0.161 | | 3,043<br>3,029 | 23.6%<br>23.5% | 0.003 |

**Supplementary Table 6. Dynamic versus low inflammation in the period surrounding a psychosocial or socioeconomic stressor.** Demographic characteristics and other covariates of interest, including types of adverse events, comorbidities and hospital admissions within the last month before the record of adversity, in patients with dynamic changes in (Cohort 1) and persistent normal leukocyte counts (Cohort 2) in the period surrounding a record of a psychosocial stressor, before and after matching. SMD: standardised mean difference.

| **Cohort characteristics** | | **Cohort 1 (N = 34,249) and Cohort 2 (N = 100,061) before propensity score matching** | | | | **Cohort 1 (N = 31,447) and Cohort 2 (N = 31,447) after propensity score matching** | | | |
|---|---|---|---|---|---|---|---|---|---|
| **Demographics** | | | | | | | | | |
| Cohort | | Mean ± SD | Patients | % of Cohort | SMD | Mean ± SD | Patients | % of Cohort | SMD |
| 1<br>2 | Age at Index | 49.6 +/- 17.2<br>48.9 +/- 17.2 | 34,249<br>100,061 | 100%<br>100% | 0.037 | 49.3 +/- 17.3<br>49.2 +/- 17.2 | 31,447<br>31,447 | 100%<br>100% | 0.005 |
| 1<br>2 | Female | | 16,438<br>51,525 | 48.0%<br>51.5% | 0.070 | | 15,325<br>15,347 | 48.7%<br>48.8% | 0.001 |
| 1<br>2 | Male | | 16,080<br>42,907 | 47.0%<br>42.9% | 0.082 | | 14,530<br>14,646 | 46.2%<br>46.6% | 0.007 |
| 1<br>2 | Black or African American | | 6,690<br>24,968 | 19.5%<br>25.0% | 0.131 | | 6,339<br>6,142 | 20.2%<br>19.5% | 0.016 |
| 1<br>2 | White | | 20,255<br>53,476 | 59.1%<br>53.4% | 0.115 | | 18,385<br>18,782 | 58.5%<br>59.7% | 0.026 |
| 1<br>2 | Asian | | 883<br>2,308 | 2.6%<br>2.3% | 0.018 | | 742<br>692 | 2.4%<br>2.2% | 0.011 |
| 1<br>2 | Native Hawaiian or Other Pacific Islander | | 502<br>838 | 1.5%<br>0.8% | 0.059 | | 405<br>392 | 1.3%<br>1.2% | 0.004 |
| 1<br>2 | American Indian or Alaska Native | | 429<br>954 | 1.3%<br>1.0% | 0.029 | | 394<br>356 | 1.3%<br>1.1% | 0.011 |
| 1<br>2 | Other Race | | 1,013<br>3,228 | 3.0%<br>3.2% | 0.015 | | 950<br>854 | 3.0%<br>2.7% | 0.018 |
| 1<br>2 | Unknown Race | | 4,477<br>14,289 | 13.1%<br>14.3% | 0.035 | | 4,232<br>4,229 | 13.5%<br>13.4% | <0.001 |
| 1<br>2 | Hispanic or Latino | | 3,317<br>9,357 | 9.7%<br>9.4% | 0.011 | | 3,046<br>2,905 | 9.7%<br>9.2% | 0.015 |
| 1<br>2 | Not Hispanic or Latino | | 21,395<br>60,226 | 62.5%<br>60.2% | 0.047 | | 19,391<br>19,455 | 61.7%<br>61.9% | 0.004 |
| 1<br>2 | Unknown Ethnicity | | 9,537<br>30,478 | 27.8%<br>30.5% | 0.058 | | 9,010<br>9,087 | 28.7%<br>28.9% | 0.005 |
| **Diagnosis** | | | | | | | | | |
| 1<br>2 | Mental and behavioral disorders due to psychoactive substance use | | 16,371<br>40,255 | 47.8%<br>40.2% | 0.153 | | 14,846<br>14,204 | 47.2%<br>45.2% | 0.041 |

| | | | | | | | |
|---|---|---|---|---|---|---|---|
| 1<br>2 | Mood [affective] disorders | 12,858<br>38,816 | 37.5%<br>38.8% | 0.026 | 11,758<br>12,862 | 37.4%<br>40.9% | 0.072 |
| 1<br>2 | Anxiety, dissociative, stress-related, somatoform and other nonpsychotic mental disorders | 12,493<br>34,922 | 36.5%<br>34.9% | 0.033 | 11,314<br>11,402 | 36.0%<br>36.3% | 0.006 |
| 1<br>2 | Schizophrenia, schizotypal, delusional, and other non-mood psychotic disorders | 2,998<br>10,791 | 8.8%<br>10.8% | 0.068 | 2,807<br>3,182 | 8.9%<br>10.1% | 0.041 |
| 1<br>2 | Disorders of adult personality and behavior | 1,525<br>4,842 | 4.5%<br>4.8% | 0.018 | 1,410<br>1,591 | 4.5%<br>5.1% | 0.027 |
| 1<br>2 | Intellectual Disabilities | 340<br>1,094 | 1.0%<br>1.1% | 0.010 | 303<br>368 | 1.0%<br>1.2% | 0.020 |
| 1<br>2 | Pervasive and specific developmental disorders | 302<br>1,067 | 0.9%<br>1.1% | 0.019 | 276<br>359 | 0.9%<br>1.1% | 0.026 |
| 1<br>2 | Polyneuropathies and other disorders of the peripheral nervous system | 1,605<br>3,558 | 4.7%<br>3.6% | 0.057 | 1,396<br>1,457 | 4.4%<br>4.6% | 0.009 |
| 1<br>2 | Nerve, nerve root and plexus disorders | 815<br>1,781 | 2.4%<br>1.8% | 0.042 | 716<br>624 | 2.3%<br>2.0% | 0.020 |
| 1<br>2 | Extrapyramidal and movement disorders | 1,222<br>2,876 | 3.6%<br>2.9% | 0.039 | 1,068<br>1,139 | 3.4%<br>3.6% | 0.012 |
| 1<br>2 | Inflammatory diseases of the central nervous system | 489<br>420 | 1.4%<br>0.4% | 0.106 | 384<br>252 | 1.2%<br>0.8% | 0.042 |
| 1<br>2 | Diseases of myoneural junction and muscle | 335<br>442 | 1.0%<br>0.4% | 0.064 | 279<br>198 | 0.9%<br>0.6% | 0.030 |
| 1<br>2 | Demyelinating diseases of the central nervous system | 294<br>642 | 0.9%<br>0.6% | 0.025 | 264<br>204 | 0.8%<br>0.6% | 0.022 |
| 1<br>2 | Neoplasms | 6,093<br>13,291 | 17.8%<br>13.3% | 0.125 | 5,546<br>4,468 | 17.6%<br>14.2% | 0.094 |
| 1<br>2 | Type 1 diabetes mellitus | 1,132<br>1,860 | 3.3%<br>1.9% | 0.091 | 988<br>739 | 3.1%<br>2.3% | 0.048 |
| 1<br>2 | Type 2 diabetes mellitus | 9,299<br>20,475 | 27.2%<br>20.5% | 0.158 | 8,259<br>7,352 | 26.3%<br>23.4% | 0.067 |
| 1<br>2 | Overweight and obesity | 7,187<br>14,007 | 21.0%<br>14.0% | 0.185 | 6,449<br>5,118 | 20.5%<br>16.3% | 0.109 |
| 1<br>2 | Hypertensive diseases | 17,005<br>40,125 | 49.7%<br>40.1% | 0.193 | 15,167<br>14,121 | 48.2%<br>44.9% | 0.067 |
| 1<br>2 | Other forms of heart disease | 11,905<br>21,445 | 34.8%<br>21.4% | 0.300 | 10,175<br>8,922 | 32.4%<br>28.4% | 0.087 |
| 1<br>2 | Ischemic heart diseases | 6,649<br>12,270 | 19.4%<br>12.3% | 0.197 | 5,674<br>4,910 | 18.0%<br>15.6% | 0.065 |

| | | | | | | | | |
|---|---|---|---|---|---|---|---|---|
| 1<br>2 | Diseases of the blood and blood-forming organs and certain disorders involving the immune mechanism | 16,807<br>25,737 | 49.1%<br>25.7% | 0.497 | | 14,256<br>14,123 | 45.3%<br>44.9% | 0.008 |
| 1<br>2 | Certain infectious and parasitic diseases | 13,501<br>20,040 | 39.4%<br>20.0% | 0.434 | | 11,357<br>11,196 | 36.1%<br>35.6% | 0.011 |
| 1<br>2 | Systemic connective tissue disorders | 710<br>1,950 | 2.1%<br>1.9% | 0.009 | | 634<br>511 | 2.0%<br>1.6% | 0.029 |
| 1<br>2 | Inflammatory polyarthropathies | 2,064<br>4,788 | 6.0%<br>4.8% | 0.055 | | 1,806<br>1,647 | 5.7%<br>5.2% | 0.022 |
| 1<br>2 | Diseases of the musculoskeletal system and connective tissue | 16,101<br>37,899 | 47.0%<br>37.9% | 0.186 | | 14,375<br>12,872 | 45.7%<br>40.9% | 0.097 |
| 1<br>2 | Certain disorders involving the immune mechanism | 1,132<br>2,365 | 3.3%<br>2.4% | 0.057 | | 1,029<br>907 | 3.3%<br>2.9% | 0.022 |
| 1<br>2 | Malignant neoplasms of lymphoid, hematopoietic and related tissue | 883<br>1,989 | 2.6%<br>2.0% | 0.040 | | 807<br>706 | 2.6%<br>2.2% | 0.021 |
| 1<br>2 | Nicotine dependence | 12,001<br>26,809 | 35.0%<br>26.8% | 0.179 | | 10,926<br>9,517 | 34.7%<br>30.3% | 0.096 |
| 1<br>2 | Encounter for examination and observation following transport accident | 38<br>52 | 0.1%<br>0.1% | 0.021 | | 31<br>13 | 0.1%<br>0.0% | 0.022 |
| 1<br>2 | Encounter for examination and observation following work accident | 10<br>10 | 0.0%<br>0.0% | 0.014 | | 10<br>10 | 0.0%<br>0.0% | <0.001 |
| 1<br>2 | Encounter for examination and observation following other accident | 205<br>421 | 0.6%<br>0.4% | 0.025 | | 181<br>163 | 0.6%<br>0.5% | 0.008 |
| 1<br>2 | Encounter for examination and observation following alleged rape | 11<br>37 | 0.0%<br>0.0% | 0.003 | | 10<br>16 | 0.0%<br>0.1% | 0.009 |
| 1<br>2 | Encounter for examination and observation following alleged physical abuse | 10<br>27 | 0.0%<br>0.0% | 0.001 | | 10<br>10 | 0.0%<br>0.0% | <0.001 |
| 1<br>2 | Problems related to social environment | 6,293<br>16,737 | 18.4%<br>16.7% | 0.043 | | 5,762<br>5,718 | 18.3%<br>18.2% | 0.004 |
| 1<br>2 | Other problems related to primary support group, including family circumstances | 8,403<br>28,107 | 24.5%<br>28.1% | 0.081 | | 7,707<br>7,413 | 24.5%<br>23.6% | 0.022 |
| 1<br>2 | Problems related to housing and economic circumstances | 17,703<br>49,946 | 51.7%<br>49.9% | 0.035 | | 16,229<br>16,543 | 51.6%<br>52.6% | 0.020 |

| | | | | | | | |
|---|---|---|---|---|---|---|---|
| 1<br>2 | Problems related to employment and unemployment | 4,739<br>14,258 | 13.8%<br>14.2% | 0.012 | 4,400<br>4,140 | 14.0%<br>13.2% | 0.024 |
| 1<br>2 | Major depressive disorder, recurrent | 2,390<br>9,708 | 7.0%<br>9.7% | 0.099 | 2,224<br>3,018 | 7.1%<br>9.6% | 0.091 |
| **Procedure** | | | | | | | |
| 1<br>2 | Hospital Inpatient and Observation Care Services | 14,845<br>21,547 | 43.3%<br>21.5% | 0.479 | 12,434<br>12,607 | 39.5%<br>40.1% | 0.011 |
| 1<br>2 | Critical Care Services | 5,631<br>3,576 | 16.4%<br>3.6% | 0.439 | 3,321<br>3,148 | 10.6%<br>10.0% | 0.018 |

**Supplementary Table 7. Dynamic versus high inflammation in the period surrounding a psychosocial or socioeconomic stressor.** Demographic characteristics and other covariates of interest, including types of adverse events, comorbidities and hospital admissions within the last month before the record of adversity, in patients with dynamic changes in (Cohort 1) and persistent high leukocyte counts (Cohort 2) in the period surrounding a record of a psychosocial stressor, before and after matching. SMD: standardised mean difference.

| **Cohort characteristics** | | **Cohort 1 (N = 34,249) and Cohort 2 (N = 31,118) before propensity score matching** | | | | **Cohort 1 (N = 30,842) and Cohort 2 (N = 30,842) after propensity score matching** | | | |
|---|---|---|---|---|---|---|---|---|---|
| **Demographics** | | | | | | | | | |
| Cohort | | Mean ± SD | Patients | % of Cohort | SMD | Mean ± SD | Patients | % of Cohort | SMD |
| 1<br>2 | Age at Index | 49.6 +/- 17.2<br>49.2 +/- 17.2 | 34,249<br>31,118 | 100%<br>100% | 0.021 | 49.3 +/- 17.2<br>49.2 +/- 17.2 | 30,842<br>30,842 | 100%<br>100% | 0.002 |
| 1<br>2 | Female | | 16,438<br>15,173 | 48.0%<br>48.8% | 0.015 | | 15,016<br>15,023 | 48.7%<br>48.7% | <0.001 |
| 1<br>2 | Male | | 16,080<br>14,330 | 47.0%<br>46.1% | 0.018 | | 14,234<br>14,228 | 46.2%<br>46.1% | <0.001 |
| 1<br>2 | Black or African American | | 6,690<br>6,132 | 19.5%<br>19.7% | 0.004 | | 5,996<br>6,078 | 19.4%<br>19.7% | 0.007 |
| 1<br>2 | White | | 20,255<br>18,594 | 59.1%<br>59.8% | 0.012 | | 18,628<br>18,419 | 60.4%<br>59.7% | 0.014 |
| 1<br>2 | Asian | | 883<br>769 | 2.6%<br>2.5% | 0.007 | | 750<br>768 | 2.4%<br>2.5% | 0.004 |
| 1<br>2 | Native Hawaiian or Other Pacific Islander | | 502<br>474 | 1.5%<br>1.5% | 0.005 | | 462<br>468 | 1.5%<br>1.5% | 0.002 |
| 1<br>2 | American Indian or Alaska Native | | 429<br>330 | 1.3%<br>1.1% | 0.018 | | 317<br>329 | 1.0%<br>1.1% | 0.004 |
| 1<br>2 | Other Race | | 1,013<br>923 | 3.0%<br>3.0% | <0.001 | | 892<br>913 | 2.9%<br>3.0% | 0.004 |
| 1<br>2 | Unknown Race | | 4,477<br>3,896 | 13.1%<br>12.5% | 0.017 | | 3,797<br>3,867 | 12.3%<br>12.5% | 0.007 |
| 1<br>2 | Hispanic or Latino | | 3,317<br>2,869 | 9.7%<br>9.2% | 0.016 | | 2,805<br>2,859 | 9.1%<br>9.3% | 0.006 |
| 1<br>2 | Not Hispanic or Latino | | 21,395<br>19,638 | 62.5%<br>63.1% | 0.013 | | 19,520<br>19,443 | 63.3%<br>63.0% | 0.005 |
| 1<br>2 | Unknown Ethnicity | | 9,537<br>8,611 | 27.8%<br>27.7% | 0.004 | | 8,517<br>8,540 | 27.6%<br>27.7% | 0.002 |
| **Diagnosis** | | | | | | | | | |
| 1<br>2 | Mental and behavioral disorders due to psychoactive substance use | | 16,371<br>14,830 | 47.8%<br>47.7% | 0.003 | | 14,723<br>14,712 | 47.7%<br>47.7% | 0.001 |

| | | | | | | | |
|---|---|---|---|---|---|---|---|
| 1<br>2 | Mood [affective] disorders | 12,858<br>11,625 | 37.5%<br>37.4% | 0.004 | 11,708<br>11,511 | 38.0%<br>37.3% | 0.013 |
| 1<br>2 | Anxiety, dissociative, stress-related, somatoform and other nonpsychotic mental disorders | 12,493<br>11,528 | 36.5%<br>37.0% | 0.012 | 11,429<br>11,398 | 37.1%<br>37.0% | 0.002 |
| 1<br>2 | Schizophrenia, schizotypal, delusional, and other non-mood psychotic disorders | 2,998<br>2,520 | 8.8%<br>8.1% | 0.024 | 2,667<br>2,502 | 8.6%<br>8.1% | 0.019 |
| 1<br>2 | Disorders of adult personality and behavior | 1,525<br>1,296 | 4.5%<br>4.2% | 0.014 | 1,388<br>1,286 | 4.5%<br>4.2% | 0.016 |
| 1<br>2 | Intellectual Disabilities | 340<br>251 | 1.0%<br>0.8% | 0.020 | 304<br>251 | 1.0%<br>0.8% | 0.018 |
| 1<br>2 | Pervasive and specific developmental disorders | 302<br>246 | 0.9%<br>0.8% | 0.010 | 275<br>243 | 0.9%<br>0.8% | 0.011 |
| 1<br>2 | Polyneuropathies and other disorders of the peripheral nervous system | 1,605<br>1,545 | 4.7%<br>5.0% | 0.013 | 1,471<br>1,522 | 4.8%<br>4.9% | 0.008 |
| 1<br>2 | Nerve, nerve root and plexus disorders | 815<br>743 | 2.4%<br>2.4% | 0.001 | 742<br>732 | 2.4%<br>2.4% | 0.002 |
| 1<br>2 | Extrapyramidal and movement disorders | 1,222<br>1,122 | 3.6%<br>3.6% | 0.002 | 1,127<br>1,109 | 3.7%<br>3.6% | 0.003 |
| 1<br>2 | Inflammatory diseases of the central nervous system | 489<br>435 | 1.4%<br>1.4% | 0.003 | 459<br>427 | 1.5%<br>1.4% | 0.009 |
| 1<br>2 | Diseases of myoneural junction and muscle | 335<br>476 | 1.0%<br>1.5% | 0.050 | 318<br>469 | 1.0%<br>1.5% | 0.044 |
| 1<br>2 | Demyelinating diseases of the central nervous system | 294<br>264 | 0.9%<br>0.8% | 0.001 | 272<br>262 | 0.9%<br>0.8% | 0.003 |
| 1<br>2 | Neoplasms | 6,093<br>6,092 | 17.8%<br>19.6% | 0.046 | 5,605<br>5,980 | 18.2%<br>19.4% | 0.031 |
| 1<br>2 | Type 1 diabetes mellitus | 1,132<br>1,143 | 3.3%<br>3.7% | 0.020 | 1,032<br>1,134 | 3.3%<br>3.7% | 0.018 |
| 1<br>2 | Type 2 diabetes mellitus | 9,299<br>9,243 | 27.2%<br>29.7% | 0.057 | 8,367<br>9,159 | 27.1%<br>29.7% | 0.057 |
| 1<br>2 | Overweight and obesity | 7,187<br>6,973 | 21.0%<br>22.4% | 0.035 | 6,549<br>6,901 | 21.2%<br>22.4% | 0.028 |
| 1<br>2 | Hypertensive diseases | 17,005<br>16,135 | 49.7%<br>51.9% | 0.044 | 15,320<br>16,001 | 49.7%<br>51.9% | 0.044 |
| 1<br>2 | Other forms of heart disease | 11,905<br>11,378 | 34.8%<br>36.6% | 0.038 | 10,832<br>11,270 | 35.1%<br>36.5% | 0.030 |
| 1<br>2 | Ischemic heart diseases | 6,649<br>6,317 | 19.4%<br>20.3% | 0.022 | 6,002<br>6,263 | 19.5%<br>20.3% | 0.021 |

| | | | | | | | |
|---|---|---|---|---|---|---|---|
| 1<br>2 | Diseases of the blood and blood-forming organs and certain disorders involving the immune mechanism | 16,807<br>16,334 | 49.1%<br>52.5% | 0.068 | 16,121<br>16,075 | 52.3%<br>52.1% | 0.003 |
| 1<br>2 | Certain infectious and parasitic diseases | 13,501<br>12,782 | 39.4%<br>41.1% | 0.034 | 12,548<br>12,593 | 40.7%<br>40.8% | 0.003 |
| 1<br>2 | Systemic connective tissue disorders | 710<br>731 | 2.1%<br>2.3% | 0.019 | 685<br>703 | 2.2%<br>2.3% | 0.004 |
| 1<br>2 | Inflammatory polyarthropathies | 2,064<br>1,930 | 6.0%<br>6.2% | 0.007 | 1,883<br>1,895 | 6.1%<br>6.1% | 0.002 |
| 1<br>2 | Diseases of the musculoskeletal system and connective tissue | 16,101<br>14,480 | 47.0%<br>46.5% | 0.010 | 14,627<br>14,339 | 47.4%<br>46.5% | 0.019 |
| 1<br>2 | Certain disorders involving the immune mechanism | 1,132<br>1,195 | 3.3%<br>3.8% | 0.029 | 1,109<br>1,143 | 3.6%<br>3.7% | 0.006 |
| 1<br>2 | Malignant neoplasms of lymphoid, hematopoietic and related tissue | 883<br>1,011 | 2.6%<br>3.2% | 0.040 | 874<br>928 | 2.8%<br>3.0% | 0.010 |
| 1<br>2 | Nicotine dependence | 12,001<br>11,128 | 35.0%<br>35.8% | 0.015 | 10,821<br>11,043 | 35.1%<br>35.8% | 0.015 |
| 1<br>2 | Encounter for examination and observation following transport accident | 38<br>25 | 0.1%<br>0.1% | 0.010 | 37<br>24 | 0.1%<br>0.1% | 0.013 |
| 1<br>2 | Encounter for examination and observation following work accident | 10<br>10 | 0.0%<br>0.0% | 0.002 | 10<br>10 | 0.0%<br>0.0% | <0.001 |
| 1<br>2 | Encounter for examination and observation following other accident | 205<br>149 | 0.6%<br>0.5% | 0.016 | 188<br>148 | 0.6%<br>0.5% | 0.018 |
| 1<br>2 | Encounter for examination and observation following alleged rape | 11<br>10 | 0.0%<br>0.0% | <0.001 | 10<br>10 | 0.0%<br>0.0% | <0.001 |
| 1<br>2 | Encounter for examination and observation following alleged physical abuse | 10<br>10 | 0.0%<br>0.0% | 0.002 | 10<br>10 | 0.0%<br>0.0% | <0.001 |
| 1<br>2 | Problems related to social environment | 6,293<br>5,899 | 18.4%<br>19.0% | 0.015 | 5,824<br>5,821 | 18.9%<br>18.9% | <0.001 |
| 1<br>2 | Other problems related to primary support group, including family circumstances | 8,403<br>7,869 | 24.5%<br>25.3% | 0.017 | 7,669<br>7,774 | 24.9%<br>25.2% | 0.008 |
| 1<br>2 | Problems related to housing and economic circumstances | 17,703<br>15,824 | 51.7%<br>50.9% | 0.017 | 15,718<br>15,716 | 51.0%<br>51.0% | <0.001 |

| | | | | | | | | |
|---|---|---|---|---|---|---|---|---|
| 1<br>2 | Problems related to employment and unemployment | 4,739<br>4,285 | 13.8%<br>13.8% | 0.002 | | 4,251<br>4,250 | 13.8%<br>13.8% | <0.001 |
| 1<br>2 | Major depressive disorder, recurrent | 2,390<br>2,051 | 7.0%<br>6.6% | 0.015 | | 2,145<br>2,038 | 7.0%<br>6.6% | 0.014 |
| **Procedure** | | | | | | | | |
| 1<br>2 | Hospital Inpatient and Observation Care Services | 14,845<br>13,558 | 43.3%<br>43.6% | 0.005 | | 13,446<br>13,425 | 43.6%<br>43.5% | 0.001 |
| 1<br>2 | Critical Care Services | 5,631<br>5,017 | 16.4%<br>16.1% | 0.009 | | 4,947<br>4,983 | 16.0%<br>16.2% | 0.003 |

**Supplementary Table 8. Incidence of a negative control outcome within one week to two years after the record of a stressor in matched cohorts with mild leukocytosis and normal leukocyte counts.** Numbers of cases (%) with an outcome are provided, along with the odds ratio (OR) and *p* values (*p*<0.05 highlighted in bold).

| Outcome | N (%) with mild leukocytosis | N (%) with normal leukocytes | OR (95% CI) | *p* |
|---|---|---|---|---|
| *Adverse event* | N=37039 | N=37039 | | |
| Hallux valgus | 105 (0.3%) | 108 (0.3%) | 0.971 (0.742-1.271) | 0.832 |
| Lipoma | 137 (0.4%) | 131 (0.4%) | 1.044 (0.821-1.326) | 0.728 |
| Ingrown nail | 133 (0.4%) | 164 (0.4%) | 0.808 (0.643-1.016) | 0.068 |
| *Psychosocial stressor* | N=83128 | N=83128 | | |
| Hallux valgus | 216 (0.3%) | 255 (0.3%) | 0.845 (0.705-1.013) | 0.069 |
| Lipoma | 319 (0.4%) | 348 (0.4%) | 0.916 (0.786-1.066) | 0.257 |
| Ingrown nail | 411 (0.5%) | 397 (0.5%) | 1.033 (0.900-1.187) | 0.641 |

**Supplementary Table 9. Incidence of a new outcome within one week to two years after the record of an adverse event in matched cohorts with leukocytosis only surrounding adversity versus persistent normal or high leukocyte counts after excluding cases with corticosteroid treatment.** Numbers of cases (%) with an outcome are provided, along with the odds ratio (OR) and *p* values (*p*<0.05 highlighted in bold).

| | DYNAMIC vs. LOW INFLAMMATION | | | | DYNAMIC vs. HIGH INFLAMMATION | | | |
|---|---|---|---|---|---|---|---|---|
| **Outcome** | **N (%) with leukocyte count change** | **N (%) with normal leukocytes** | **OR (95% CI)** | ***p*** | **N (%) with leukocyte count change** | **N (%) with high leukocytes** | **OR (95% CI)** | ***p*** |
| *Adverse event* | N=12184 | N=12184 | | | N=12849 | N=12849 | | |
| Hallux valgus | 54 (0.5%) | 67 (0.6%) | 0.804 (0.561-1.151) | 0.233 | 54 (0.4%) | 58 (0.5%) | 0.932 (0.643-1.351) | 0.709 |
| Lipoma | 83 (0.7%) | 71 (0.6%) | 1.166 (0.848-1.602) | 0.344 | 82 (0.6%) | 77 (0.6%) | 1.067 (0.781-1.458) | 0.684 |
| Ingrown nail | 71 (0.6%) | 81 (0.7%) | 0.872 (0.634-1.201) | 0.402 | 77 (0.6%) | 76 (0.6%) | 1.017 (0.740-1.398) | 0.917 |
| *Psychosocial stressor* | N=30603 | N=30603 | | | N=29621 | N=29621 | | |
| Hallux valgus | 116 (0.4%) | 145 (0.5%) | 0.798 (0.625-1.020) | 0.072 | 112 (0.4%) | 88 (0.3%) | 1.277 (0.965-1.689) | 0.086 |
| Lipoma | 168 (0.6%) | 177 (0.6%) | 0.946 (0.767-1.168) | 0.610 | 169 (0.6%) | 170 (0.6%) | 0.994 (0.803-1.231) | 0.957 |
| Ingrown nail | 217 (0.7%) | 218 (0.7%) | 0.992 (0.822-1.198) | 0.934 | 202 (0.7%) | 229 (0.8%) | 0.881 (0.729-1.065) | 0.191 |

**Supplementary Table 10. Incidence of a new outcome within one week to two years after the record of an adverse event in matched cohorts with mild leukocytosis and normal leukocyte counts after excluding cases with corticosteroid treatment.** Numbers of cases (%) with an outcome are provided, along with the odds ratio (OR) and *p* values. *p* values significant after FDR correction for 17 variables are highlighted in bold.

| Outcome | N (%) with mild leukocytosis | N (%) with normal leukocytes | OR (95% CI) | *p* | *p* (FDR) |
|---|---|---|---|---|---|
| | N=27947 | N=27947 | | | |
| *Mental health/interface conditions* | | | | | |
| Anxiety disorders | 1361 (6.1%) | 1452 (6.7%) | 0.898 (0.832-0.970) | 0.006 | **0.020** |
| Depression/depressive episode | 1236 (5.6%) | 1297 (6.0%) | 0.916 (0.846-0.993) | 0.033 | 0.056 |
| PTSD | 460 (1.7%) | 483 (1.8%) | 0.945 (0.833-1.073) | 0.381 | 0.432 |
| FND | 84 (0.3%) | 91 (0.3%) | 0.920 (0.684-1.239) | 0.583 | 0.619 |
| Somatoform disorders | 91 (0.3%) | 115 (0.4%) | 0.789 (0.599-1.039) | 0.090 | 0.128 |
| Sleep disorders | 285 (1.0%) | 338 (1.2%) | 0.837 (0.715-0.981) | 0.028 | 0.053 |
| Cognitive symptoms | 993 (4.1%) | 1103 (4.6%) | 0.885 (0.811-0.966) | 0.006 | **0.020** |
| *Pain, somatic symptoms, fatigue* | | | | | |
| Headache | 1111 (5.1%) | 1195 (5.8%) | 0.872 (0.802-0.948) | 0.001 | **0.009** |
| Unspecified chest pain | 1021 (4.8%) | 1089 (5.3%) | 0.900 (0.825-0.983) | 0.019 | **0.046** |
| Unspecified abdominal pain | 995 (4.6%) | 1128 (5.3%) | 0.850 (0.779-0.928) | <0.001 | **0.009** |
| Low back pain | 1118 (5.2%) | 1150 (5.7%) | 0.931 (0.856-1.013) | 0.099 | 0.129 |
| Fibromyalgia | 134 (0.5%) | 128 (0.5%) | 1.042 (0.817-1.328) | 0.740 | 0.740 |
| Breathing abnormalities | 1406 (6.4%) | 1464 (6.8%) | 0.932 (0.864-1.005) | 0.069 | 0.107 |
| Palpitations | 388 (1.5%) | 471 (1.8%) | 0.812 (0.709-0.930) | 0.003 | **0.017** |
| Irritable bowel syndrome | 106 (0.4%) | 123 (0.5%) | 0.859 (0.662-1.114) | 0.251 | 0.305 |
| Pruritus | 227 (0.8%) | 275 (1.0%) | 0.818 (0.686-0.976) | 0.026 | 0.053 |
| Malaise and fatigue | 1388 (6.0%) | 1453 (6.5%) | 0.913 (0.846-0.985) | 0.019 | **0.046** |

**Supplementary Table 11. Incidence of a new outcome within one month to two years after the record of a psychosocial or socioeconomic stressor in matched cohorts with mild leukocytosis and normal leukocyte counts after excluding cases with corticosteroid treatment.** Numbers of cases (%) with an outcome are provided, along with the odds ratio (OR) and *p* values. *p* values significant after FDR correction for 17 variables are highlighted in bold.

| **Outcome** | **N (%) with mild leukocytosis** | **N (%) with normal leukocytes** | **OR (95% CI)** | ***p*** | ***p* (FDR)** |
|---|---|---|---|---|---|
| | N=52604 | N=52604 | | | |
| *Mental health/interface conditions* | | | | | |
| Anxiety disorders | 2861 (9.2%) | 3112 (10.2%) | 0.888 (0.842-0.937) | <0.001 | **0.009** |
| Depression/depressive episode | 2557 (9.0%) | 2646 (9.5%) | 0.939 (0.887-0.994) | 0.029 | 0.123 |
| Adjustment disorders | 1072 (2.3%) | 1155 (2.4%) | 0.925 (0.850-1.006) | 0.068 | 0.165 |
| FND | 197 (0.4%) | 173 (0.3%) | 1.138 (0.927-1.396) | 0.216 | 0.322 |
| Somatoform disorders | 190 (0.4%) | 210 (0.4%) | 0.904 (0.743-1.101) | 0.315 | 0.412 |
| Sleep disorders | 692 (1.4%) | 760 (1.5%) | 0.907 (0.817-1.006) | 0.064 | 0.165 |
| Cognitive symptoms | 1882 (4.2%) | 1947 (4.2%) | 0.986 (0.924-1.052) | 0.676 | 0.718 |
| *Pain, somatic symptoms, fatigue* | | | | | |
| Headache | 2593 (6.1%) | 2583 (6.1%) | 1.002 (0.947-1.060) | 0.068 | 0.165 |
| Unspecified chest pain | 2669 (6.7%) | 2668 (6.7%) | 1.010 (0.955-1.067) | 0.728 | 0.728 |
| Unspecified abdominal pain | 2542 (6.2%) | 2636 (6.3%) | 0.982 (0.929-1.039) | 0.534 | 0.648 |
| Low back pain | 2191 (5.0%) | 2280 (5.3%) | 0.964 (0.907-1.023) | 0.227 | 0.322 |
| Fibromyalgia | 330 (0.7%) | 298 (0.6%) | 1.107 (0.946-1.296) | 0.203 | 0.322 |
| Breathing abnormalities | 3158 (8.1%) | 3044 (7.6%) | 1.068 (1.014-1.125) | 0.013 | 0.074 |
| Palpitations | 943 (1.9%) | 990 (2.0%) | 0.943 (0.861-1.031) | 0.199 | 0.322 |
| Irritable bowel syndrome | 260 (0.5%) | 329 (0.6%) | 0.789 (0.670-0.929) | 0.004 | **0.034** |
| Pruritus | 548 (1.1%) | 561 (1.1%) | 0.974 (0.865-1.096) | 0.659 | 0.718 |
| Malaise and fatigue | 2884 (7.0%) | 2957 (7.3%) | 0.954 (0.905-1.007) | 0.086 | 0.183 |

**Supplementary Table 12. Incidence of a new outcome within one week to two years after the record of an adverse event in matched cohorts with leukocytosis only surrounding adversity versus persistent normal or high leukocyte counts after excluding cases with corticosteroid treatment.** Numbers of cases (%) with an outcome are provided, along with the odds ratio (OR) and *p* values. *p* values significant after FDR correction for 17 variables are highlighted in bold.

| | **DYNAMIC vs. LOW INFLAMMATION** (N = 6980) | | | | | **DYNAMIC vs. HIGH INFLAMMATION** (N = 6556) | | | | |
|---|---|---|---|---|---|---|---|---|---|---|
| **Outcome** | **N (%) with leukocyte count change** | **N (%) with normal leukocytes** | **OR (95% CI)** | ***p*** | ***p* (FDR)** | **N (%) with leukocyte count change** | **N (%) with high leukocytes** | **OR (95% CI)** | ***p*** | ***p* (FDR)** |
| *Mental health/interface conditions* | | | | | | | | | | |
| Anxiety disorders | 528 (11.4%) | 555 (12.8%) | 0.872 (0.768-0.990) | 0.034 | 0.064 | 496 (11.3%) | 587 (12.8%) | 0.884 (0.790-0.989) | 0.031 | 0.112 |
| Depression/depressive episode | 504 (11.1%) | 517 (12.6%) | 0.866 (0.760-0.986) | 0.030 | 0.064 | 467 (10.9%) | 546 (12.2%) | 0.879 (0.771-1.002) | 0.054 | 0.153 |
| PTSD | 167 (2.5%) | 154 (2.4%) | 1.078 (0.863-1.345) | 0.508 | 0.540 | 157 (2.6%) | 186 (3.0%) | 0.846 (0.682-1.049) | 0.127 | 0.308 |
| FND | 38 (0.6%) | 51 (0.7%) | 0.740 (0.487-1.125) | 0.158 | 0.203 | 34 (0.5%) | 45 (0.7%) | 0.756 (0.484-1.182) | 0.219 | 0.421 |
| Somatoform disorders | 47 (0.7%) | 61 (0.9%) | 0.765 (0.522-1.120) | 0.167 | 0.203 | 41 (0.6%) | 45 (0.7%) | 0.915 (0.598-1.398) | 0.680 | 0.826 |
| Sleep disorders | 133 (2.0%) | 165 (2.5%) | 0.798 (0.634-1.006) | 0.055 | 0.085 | 118 (1.9%) | 122 (1.9%) | 0.977 (0.757-1.261) | 0.857 | 0.971 |
| Cognitive symptoms | 500 (9.4%) | 615 (11.7%) | 0.781 (0.690-0.885) | <0.001 | **0.003** | 471 (9.5%) | 623 (12.7%) | 0.720 (0.634-0.817) | <0.001 | **0.003** |
| *Pain, somatic symptoms, fatigue* | | | | | | | | | | |
| Headache | 451 (9.0%) | 535 (11.6%) | 0.752 (0.659-0.858) | <0.001 | **0.003** | 430 (9.1%) | 438 (8.8%) | 1.038 (0.903-1.194) | 0.596 | 0.779 |
| Unspecified chest pain | 431 (9.7%) | 506 (12.3%) | 0.765 (0.668-0.877) | <0.001 | **0.003** | 404 (9.6%) | 484 (11.0%) | 0.860 (0.748-0.988) | 0.033 | 0.112 |
| Unspecified abdominal pain | 432 (9.0%) | 485 (10.9%) | 0.802 (0.699-0.919) | 0.001 | **0.004** | 406 (9.0%) | 525 (11.3%) | 0.776 (0.677-0.890) | <0.001 | **0.003** |
| Low back pain | 447 (9.0%) | 469 (10.0%) | 0.888 (0.775-1.018) | 0.087 | 0.123 | 414 (8.9%) | 414 (8.4%) | 1.056 (0.916-1.218) | 0.451 | 0.639 |
| Fibromyalgia | 47 (0.7%) | 52 (0.8%) | 0.888 (0.598-1.319) | 0.556 | 0.556 | 43 (0.7%) | 35 (0.6%) | 1.231 (0.787-1.925) | 0.362 | 0.559 |
| Breathing abnormalities | 582 (13.2%) | 611 (15.0%) | 0.861 (0.762-0.973) | 0.017 | **0.041** | 546 (13.2%) | 681 (16.5%) | 0.765 (0.678-0.865) | <0.001 | **0.003** |
| Palpitations | 160 (2.5%) | 211 (3.4%) | 0.729 (0.592-0.898) | 0.003 | **0.010** | 144 (2.4%) | 146 (2.4%) | 0.998 (0.791-1.260) | 0.988 | 0.988 |
| Irritable bowel syndrome | 48 (0.7%) | 58 (0.9%) | 0.815 (0.555-1.197) | 0.297 | 0.337 | 41 (0.6%) | 41 (0.6%) | 1.005 (0.651-1.552) | 0.981 | 0.988 |
| Pruritus | 106 (1.6%) | 144 (2.2%) | 0.718 (0.557-0.925) | 0.010 | **0.028** | 96 (1.5%) | 110 (1.7%) | 0.874 (0.663-1.151) | 0.337 | 0.559 |
| Malaise and fatigue | 634 (13.6%) | 638 (15.1%) | 0.883 (0.784-0.995) | 0.040 | 0.068 | 618 (14.0%) | 689 (14.8%) | 0.930 (0.827-1.045) | 0.223 | 0.421 |

**Supplementary Table 13. Incidence of a new outcome within one month to two years after the record of a psychosocial or socioeconomic stressor in matched cohorts with leukocytosis only surrounding adversity versus persistent normal or high leukocyte counts after excluding cases with corticosteroid treatment.** Numbers of cases (%) with an outcome are provided, along with the odds ratio (OR) and *p* values. *p* values significant after FDR correction for 17 variables are highlighted in bold.

| | **DYNAMIC vs. LOW INFLAMMATION** (N = 15508) | | | | | **DYNAMIC vs. HIGH INFLAMMATION** (N = 10168) | | | | |
|---|---|---|---|---|---|---|---|---|---|---|
| **Outcome** | **N (%) with leukocyte count change** | **N (%) with normal leukocytes** | **OR (95% CI)** | ***p*** | ***p* (FDR)** | **N (%) with leukocyte count change** | **N (%) with high leukocytes** | **OR (95% CI)** | ***p*** | ***p* (FDR)** |
| *Mental health/interface conditions* | | | | | | | | | | |
| Anxiety disorders | 1208 (14.2%) | 1312 (16.2%) | 0.853 (0.784-0.928) | <0.001 | **0.009** | 790 (14.5%) | 855 (15.7%) | 0.912 (0.822-1.013) | 0.087 | 0.211 |
| Depression/depressive episode | 1105 (14.3%) | 1174 (16.0%) | 0.875 (0.800-0.956) | 0.003 | **0.017** | 745 (14.9%) | 748 (15.0%) | 0.994 (0.890-1.109) | 0.910 | 0.967 |
| Adjustment disorders | 512 (3.7%) | 615 (4.5%) | 0.822 (0.729-0.927) | 0.001 | **0.009** | 353 (3.9%) | 365 (4.1%) | 0.956 (0.824-1.110) | 0.557 | 0.676 |
| FND | 87 (0.6%) | 125 (0.8%) | 0.692 (0.526-0.911) | 0.008 | **0.026** | 55 (0.6%) | 83 (0.8%) | 0.662 (0.470-0.932) | 0.017 | 0.072 |
| Somatoform disorders | 100 (0.7%) | 117 (0.8%) | 0.852 (0.652-1.113) | 0.240 | 0.291 | 66 (0.7%) | 80 (0.8%) | 0.824 (0.594-1.142) | 0.244 | 0.377 |
| Sleep disorders | 315 (2.1%) | 360 (2.4%) | 0.865 (0.742-1.007) | 0.062 | 0.105 | 213 (2.2%) | 222 (2.3%) | 0.956 (0.790-1.156) | 0.641 | 0.726 |
| Cognitive symptoms | 1062 (8.5%) | 1112 (8.8%) | 0.966 (0.884-1.054) | 0.434 | 0.461 | 679 (8.3%) | 716 (8.8%) | 0.935 (0.838-1.044) | 0.231 | 0.377 |
| *Pain, somatic symptoms, fatigue* | | | | | | | | | | |
| Headache | 1166 (10.1%) | 1242 (11.0%) | 0.904 (0.831-0.984) | 0.019 | **0.040** | 789 (10.5%) | 779 (10.5%) | 1.001 (0.902-1.112) | 0.979 | 0.979 |
| Unspecified chest pain | 1228 (12.2%) | 1330 (13.3%) | 0.902 (0.830-0.980) | 0.015 | **0.036** | 790 (12.0%) | 815 (12.7%) | 0.939 (0.845-1.042) | 0.234 | 0.377 |
| Unspecified abdominal pain | 1208 (11.4%) | 1327 (12.5%) | 0.895 (0.823-0.972) | 0.009 | **0.026** | 799 (11.6%) | 871 (13.0%) | 0.877 (0.792-0.972) | 0.012 | 0.068 |
| Low back pain | 1013 (8.5%) | 1120 (9.6%) | 0.882 (0.807-0.964) | 0.006 | **0.026** | 672 (8.7%) | 686 (9.0%) | 0.957 (0.856-1.070) | 0.441 | 0.577 |
| Fibromyalgia | 153 (1.0%) | 160 (1.1%) | 0.948 (0.758-1.184) | 0.636 | 0.636 | 95 (1.0%) | 122 (1.3%) | 0.773 (0.590-1.012) | 0.060 | 0.204 |
| Breathing abnormalities | 1382 (14.2%) | 1439 (14.7%) | 0.958 (0.884-1.037) | 0.290 | 0.329 | 914 (14.2%) | 988 (16.0%) | 0.872 (0.791-0.961) | 0.006 | 0.068 |
| Palpitations | 400 (2.8%) | 451 (3.2%) | 0.871 (0.760-0.999) | 0.048 | 0.091 | 278 (3.0%) | 315 (3.4%) | 0.885 (0.751-1.042) | 0.143 | 0.304 |
| Irritable bowel syndrome | 111 (0.7%) | 139 (0.9%) | 0.795 (0.619-1.021) | 0.072 | 0.111 | 74 (0.8%) | 97 (1.0%) | 0.762 (0.562-1.032) | 0.078 | 0.211 |
| Pruritus | 309 (2.1%) | 347 (2.4%) | 0.877 (0.751-1.024) | 0.097 | 0.137 | 202 (2.1%) | 255 (2.6%) | 0.781 (0.648-0.942) | 0.009 | 0.068 |
| Malaise and fatigue | 1411 (13.2%) | 1446 (13.9%) | 0.948 (0.876-1.026) | 0.182 | 0.238 | 939 (13.4%) | 945 (13.9%) | 0.962 (0.873-1.060) | 0.432 | 0.577 |
